# Mobile consulting (mConsulting) as an option for accessing healthcare services for communities in remote rural areas and urban slums in low- and middle- income countries: A mixed methods study

**DOI:** 10.1101/2020.11.12.20229955

**Authors:** Bronwyn Harris, Motunrayo Ajisola, Raisa Alam, Jocelyn Antsley Watkins, Theodoros N Arvanitis, Pauline Bakibinga, Beatrice Chipwaza, Nazratun Choudhury, Olufunke Fayhun, Peter Kibe, Akinyinka Omigbodun, Eme Owoaje, Senga Pemba, Rachel Potter, Narjis Rizvi, Jackie Sturt, Jonathan Cave, Romaina Iqbal, Caroline Kabaria, Albino Kalolo, Catherine Kyobutungi, Richard Lilford, Titus Mashanya, Sylvester Ndegese, Omar Rahman, Saleem Sayani, Rita Yusuf, Frances Griffiths

## Abstract

**Objective:** Remote or mobile consulting (mConsulting) is being promoted to strengthen health systems, deliver universal health coverage and facilitate safe clinical communication during COVID-19 and beyond. We explored whether mConsulting is a viable option for communities with minimal resources in low- and middle-income countries (LMICs).

**Methods:** We reviewed evidence published since 2018 about mConsulting in LMICs and undertook a scoping study (pre-COVID) in two rural settings (Pakistan, Tanzania) and five urban slums (Kenya, Nigeria, Bangladesh), using policy/document review, secondary analysis of survey data (from the urban sites), and thematic analysis of interviews/workshops with community members, healthcare workers, digital/telecommunications experts, mConsulting providers, local and national decision-makers. Project advisory groups guided the study in each country.

**Results:** We reviewed five empirical studies and seven reviews, analysed data from 5,219 urban slum households and engaged with 419 stakeholders in rural and urban sites. Regulatory frameworks are available in each country. mConsulting services are operating through provider platforms (n=5-17) and, at community-level, some direct experience of mConsulting with healthcare workers using their own phones was reported - for emergencies, advice and care follow-up. Stakeholder willingness was high, provided challenges are addressed in technology, infrastructure, data security, confidentiality, acceptability and health system integration. mConsulting can reduce affordability barriers and facilitate care-seeking practices.

**Conclusions:** There are indications of readiness for mConsulting in communities with minimal resources. However, wider system strengthening is needed to bolster referrals, specialist services, laboratories and supply-chains to fully realise the continuity of care and responsiveness that mConsulting services offer, particularly during/beyond COVID-19.

## Introduction

Globally, coronavirus disease 2019 (COVID-19) has enforced changes in health seeking behaviour and in the organisation and delivery of healthcare services. Technology is in the spotlight as the world seeks new ways to support overburdened health systems and protect healthcare workers and populations, with innovations emerging in digital communication, education and patient management solutions.^1^ To safeguard the health workforce, the World Health Organization (WHO) is encouraging adoption of remote or mobile consulting (mConsulting) as an alternative to face-to-face consultation, where possible.^2^ mConsulting involves two-way clinical consultation between a person with a perceived health need and a healthcare provider, using mobile technology (e.g., mobile phone, tablet, laptop). Examples include but not limited to: someone living with diabetes sending a text message to their doctor for dietary advice; a teenager consulting an interactive website on sexual health; and a community nurse phoning to check on a child with a fever. While there has been a rapid uptake of mobile communication technology in low- and middle-income countries (LMICs) over the last decade, women, rural residents and poorer communities have been negatively affected by inequalities in access to resources, mobile phones, the internet, data and airtime ^3-6^. We therefore consider in our mConsulting definition the use of ‘non-mobile technology’, such as a communally-shared landline/computer, where it enables remote consulting services.^7^ Additionally, we acknowledge that an intermediary may assist the user with a consultation (e.g., a relative or neighbour). However, we exclude, from our definition of mConsulting, scenarios where a health worker separately seeks advice about a patient from a colleague, in the absence of the patient.^7^

During a pandemic, a clear benefit of mConsulting is that it reduces the need for physical contact and thereby protects frontline health workers, patients and vulnerable populations. Although a rapid acceleration of mConsulting is to be expected during the COVID-19 crisis, it is not a new form of clinical communication. In the past few years, digital technology has been promoted to strengthen health systems, deliver universal health coverage and provide quality health services, particularly in LMICs^3^. Yet, little is known about mConsulting in LMIC contexts: what services exist, who is using them and why? As a digital technology solution, mConsulting may help to facilitate safe clinical communication during COVID-19 and beyond. However, is it a viable option for communities and health systems with minimal resources?

In this study, undertaken pre-COVID-19, we explore this question by reviewing current evidence for mConsulting in LMIC contexts; and engaging with people living and providing healthcare in low resource settings in Pakistan, Tanzania, Kenya, Nigeria and Bangladesh.

### Conceptual framework: access in a complex adaptive system

We draw on our conceptual framework (elaborated in Griffiths et al., 2020) for understanding mConsulting as a two-way, complex adaptive system that connects patients and healthcare providers across a digital communication platform^7^. Complex adaptive systems are self-regulating, context-bound and unpredictable. System change is generated by the extent to which interconnected elements (people, organisations and policies) adapt and learn.^8^ Because mConsulting draws together digital, social and physical worlds, in ways that disrupt conventional understandings of time and place, it ‘has the potential to precipitate nonlinear change and feedback that could result in significant change’^7^. For poor and spatially-marginalised communities with limited healthcare options, mConsulting has the potential to improve access to quality services, even though there may be unintended barriers and challenges^7^. Following Levesque et al. (2013), we define access as ‘the possibility to identify healthcare needs, to seek healthcare services, to reach the healthcare resources, to obtain or use health care services, and to actually be offered services appropriate to the needs for care”^9^. Access is generated between healthcare users and providers - here, across the digital communication platform. It is enabled or impeded by the availability, affordability and acceptability of the (mConsulting) system.^9, 10^

### Literature review of current evidence for mConsulting in LMIC settings

To place our study within the context of current evidence, we reviewed literature, published since 2018, for evidence of mConsulting in LMIC settings. This timeframe coincides with the emergence of WHO guidance on digital interventions for health systems^11^ and recognises the rapidly changing nature of digital technology. Building on our previous reviews of two-way digital clinical communication^12-18^, with added parameters for LMIC contexts, we identified seven systematic reviews^19-25^ and five empirical studies^26-30^ involving mConsulting in LMICs, as per our definition^7^ (see, Appendix 1, for our search strategy).

The seven reviews included studies from a range of LMIC countries^19-25^, while the empirical studies were from Ghana^30^, Kenya^28^, Bangladesh^26, 29^ and India^27^. mConsulting was used in maternal, newborn and child health care in two reviews^24, 25^ and four studies^27-30^. Other conditions included chronic care (n=3)^22, 23, 26^ for non-communicable disease, diabetes, tuberculosis, HIV and cancer; general health (n=2)^20, 21^; mental health for mothers living with HIV (n=1)^27^; and adolescent health (n=1)^19^.

### Acceptability of mConsulting services

The provision of personalised care^26^ by a known and trusted health service provider or someone with authority and expertise (especially if a doctor)^29^ was identified as an important factor in the acceptability of mConsulting. Services tailored to local expectations and cultural practices contributed to service use, for example, where it was acceptable for women to receive calls from healthcare providers^27^, or, where concerned family members, including mothers-in-law, were able to participate, alongside new mothers and fathers in a remote consulting programme for maternal and child health in rural Bangladesh^29^. Additional reasons for its acceptability included low cost, provider access to patients, and reduced social stigma for certain conditions, such as family planning^28^, HIV^27^ and visible mental health interventions^27^, for example mCounselling between nurses and HIV-positive women in India encouraged treatment adherence and behavioural change^27^. Receiving health counselling happened at a mutually convenient and flexible time for users and providers; it was also seen as a more acceptable way to engage than through text messaging, especially for those with limited literacy^27^.

### Affordability and availability of the service

Free-to-use services or those costed at the local rate per call were valued by patients for their affordability^26^. Patient-led or on-demand services, for example those accessed through websites or helplines, were viewed as convenient and flexible for users^20^. Because users can choose when they want to access the service, this can improve confidentiality. Such services can be used in areas of low or erratic connectivity^20^. The knowledge that a service was available 24/7, especially during emergencies, was reassuring for diabetic patients in Bangladesh (even if they did not actually use it). This service was further valued because it provided personalised care^26^.

### Changing behaviour and effects on health outcomes

Dol (2018) found that mConsulting, during the perinatal period, increased mothers’ attendance of antenatal and postnatal services^24^. Mobile communication was seen to improve maternal, newborn and child health across many LMICs^25^. Mobile consultation with providers was found to contribute to improved treatment adherence and blood glucose self-testing for diabetic patients, alongside increased patient-provider communication more generally^23^. Johnston (2018) reported mixed evidence for clinical diabetes outcomes in mHealth interventions for diabetes care that included telephone consultations; these were not always associated with improved HbA1c levels^22^. Similarly, while Aberjirinde (2018) found that although mHealth with consulting for antenatal service positively influenced patient trust in health workers and in referral recommendations^30^, it was inconclusive whether this increased demand and utilisation of the services. Additionally, they found that use of a device, for an integrated diagnostic and clinical decision support system, helped the midwife’s processes but extended the time women were at the clinic, increasing impatience^30^.

### Challenges using mConsulting

Reported challenges included the need for users and healthcare providers to adapt to using a remote service^27^; these were identified as lack of integration of the remote service with referral systems and follow-up appointments, which could reduce patient continuity of care^29^; and inadvertent disclosure of sensitive information, such as someone’s HIV status, to others^27^. Additionally, services were found to potentially contribute to inequalities in patient access. For example, a dedicated helpline in Nigeria, providing information about self-examination for oncology patients, was mainly accessed by users with higher levels of formal education^19^.

### mConsulting in communities with minimal access to healthcare in Pakistan, Tanzania, Kenya, Nigeria and Bangladesh

Contextualised within our review of the current evidence for mConsulting in LMIC contexts, we undertook a scoping study of mConsulting in communities with minimal access to healthcare in five LMIC settings.

## Methods

### Study setting: country-context

In theory, mConsulting can be provided from anywhere in the world. However, the pragmatics are likely to be shaped by the needs of particular populations and the regulatory, technological and health system contexts in which mConsulting takes place^7^. To gain analytical traction on the interaction of digital processes with context, we studied mConsulting in remote and spatially-marginalised communities in Pakistan, Tanzania, Kenya, Nigeria and Bangladesh; five lower-middle income countries facing pervasive structural barriers to growth and development, with low income levels, socio-economic inequities and wide disparities in health outcomes and access to services^31^ (Table 1).

**Table 1.**
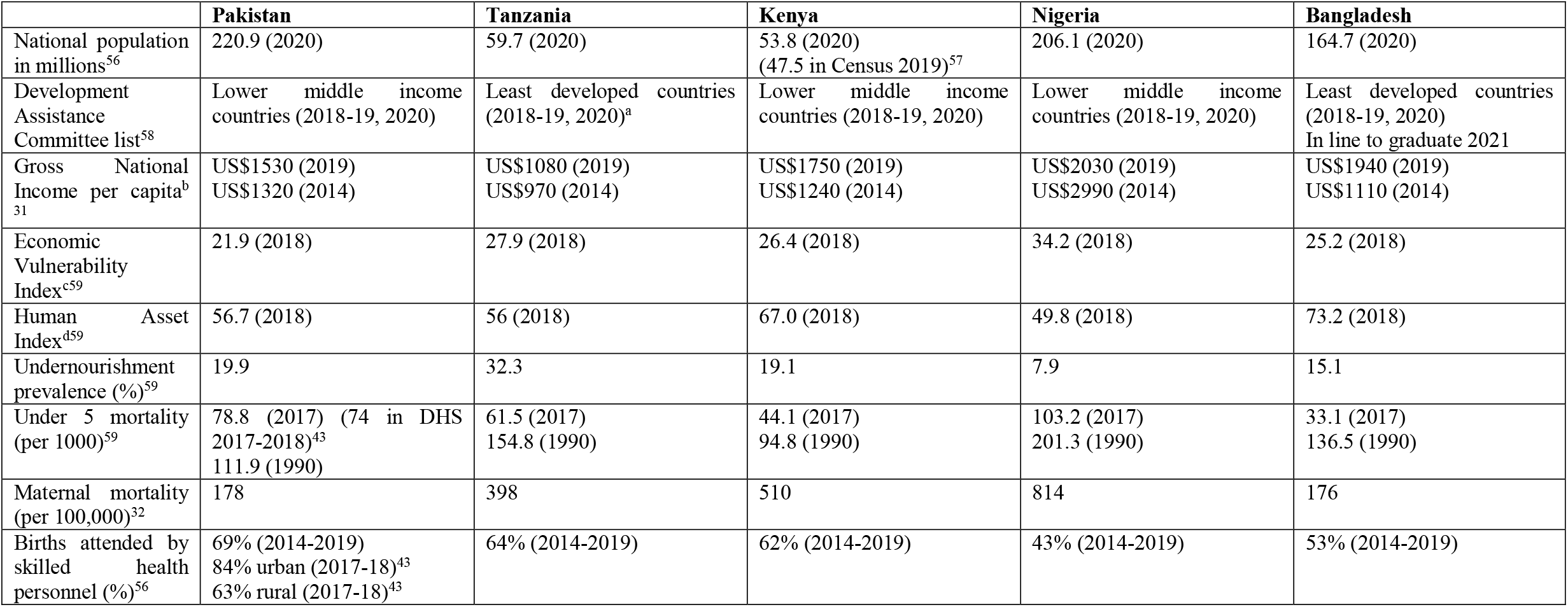
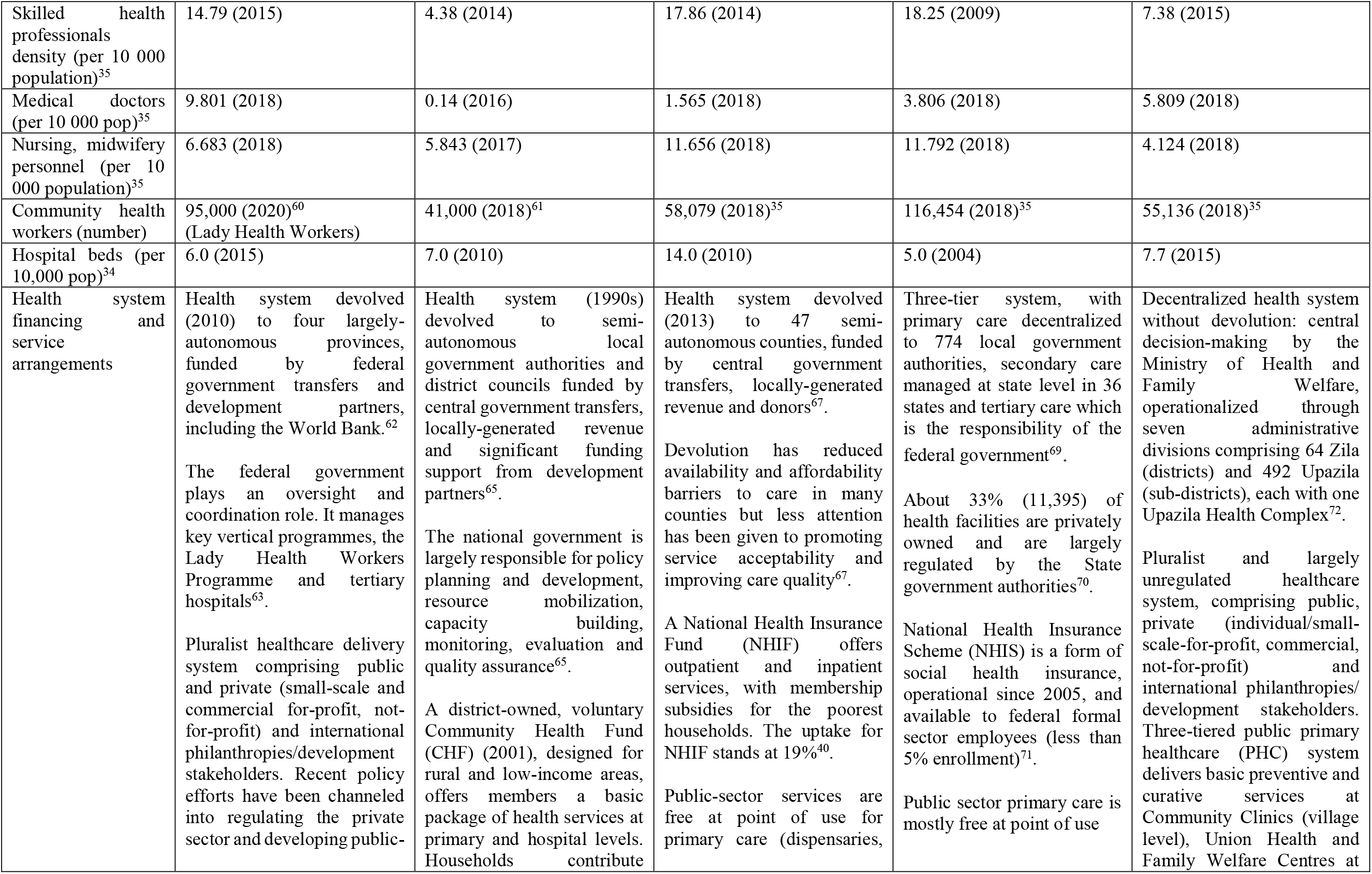
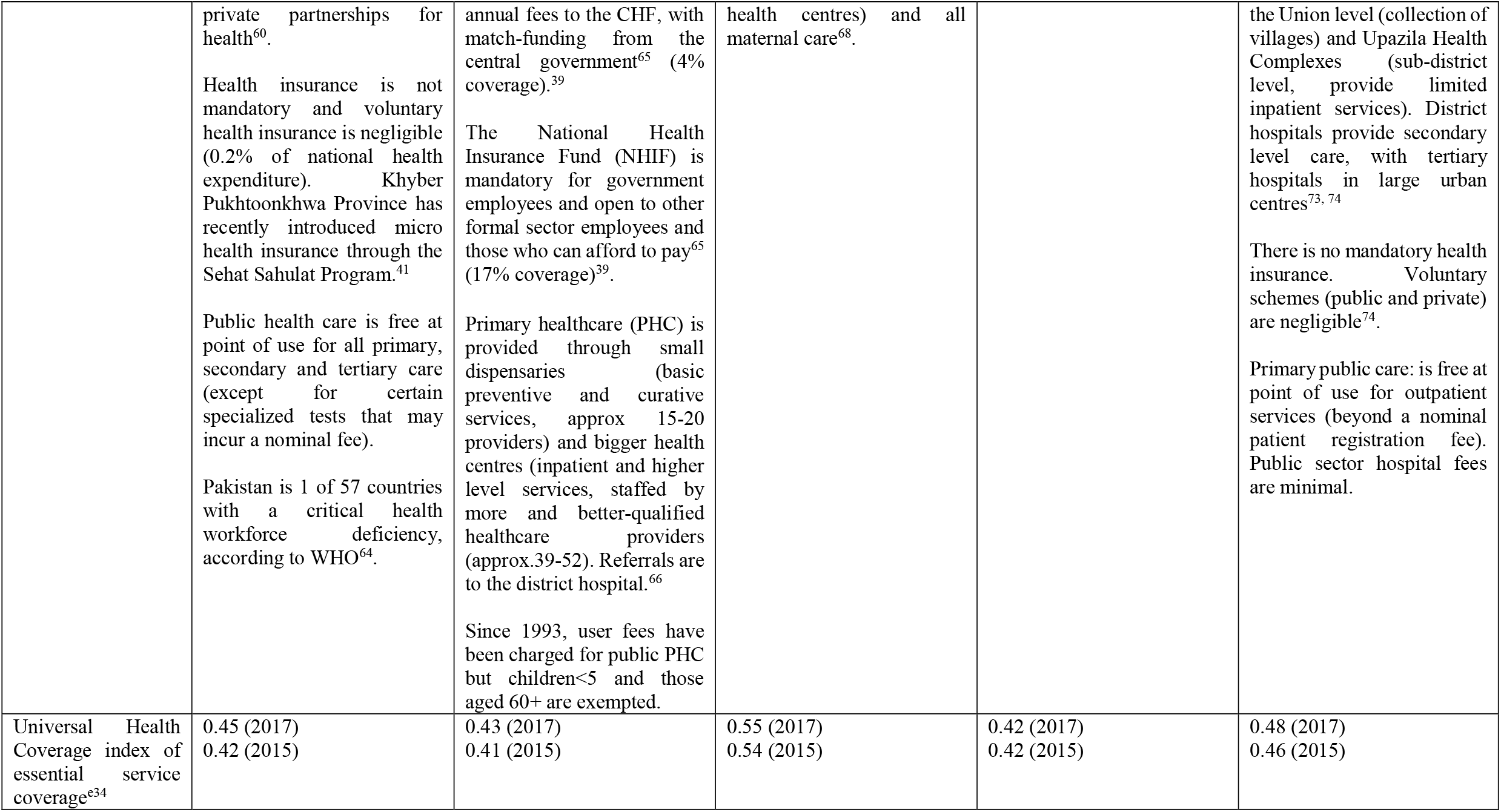
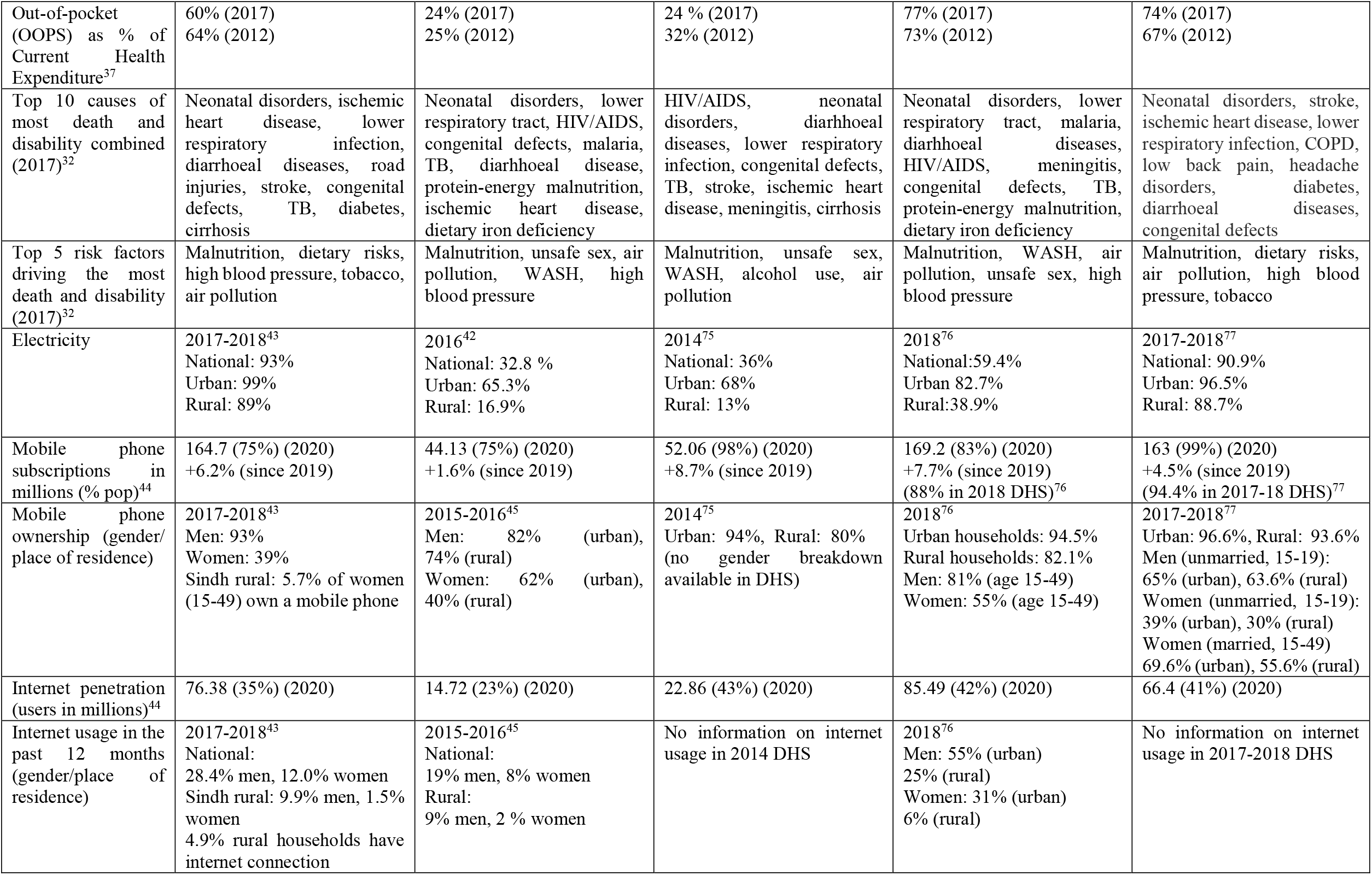
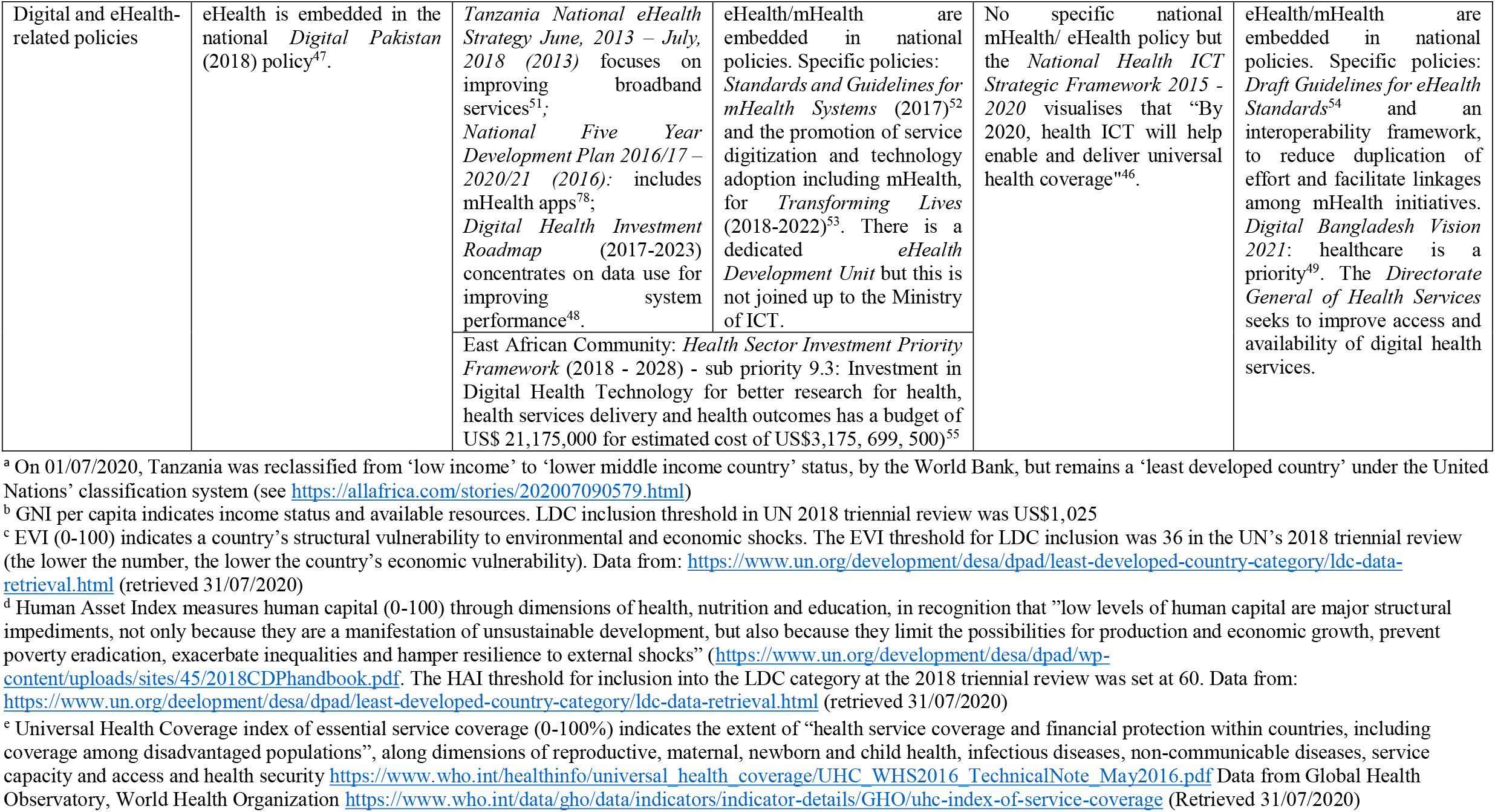
National policy and digital landscapes in Pakistan, Tanzania, Kenya, Nigeria and Bangladesh.

|  | **Pakistan** | **Tanzania** | **Kenya** | **Nigeria** | **Bangladesh** |
| --- | --- | --- | --- | --- | --- |
| National population in millions<sup>56</sup> | 220.9 (2020) | 59.7 (2020) | 53.8 (2020)<br>(47.5 in Census 2019)<sup>57</sup> | 206.1 (2020) | 164.7 (2020) |
| Development Assistance Committee list<sup>58</sup> | Lower middle income countries (2018-19, 2020) | Least developed countries (2018-19, 2020)<sup>a</sup> | Lower middle income countries (2018-19, 2020) | Lower middle income countries (2018-19, 2020) | Least developed countries (2018-19, 2020)<br>In line to graduate 2021 |
| Gross National Income per capita<sup>b</sup><br><sup>31</sup> | US\$1530 (2019)<br>US\$1320 (2014) | US\$1080 (2019)<br>US\$970 (2014) | US\$1750 (2019)<br>US\$1240 (2014) | US\$2030 (2019)<br>US\$2990 (2014) | US\$1940 (2019)<br>US\$1110 (2014) |
| Economic Vulnerability Index<sup>c59</sup> | 21.9 (2018) | 27.9 (2018) | 26.4 (2018) | 34.2 (2018) | 25.2 (2018) |
| Human Asset Index<sup>d59</sup> | 56.7 (2018) | 56 (2018) | 67.0 (2018) | 49.8 (2018) | 73.2 (2018) |
| Undernourishment prevalence (%)<sup>59</sup> | 19.9 | 32.3 | 19.1 | 7.9 | 15.1 |
| Under 5 mortality (per 1000)<sup>59</sup> | 78.8 (2017) (74 in DHS 2017-2018)<sup>43</sup><br>111.9 (1990) | 61.5 (2017)<br>154.8 (1990) | 44.1 (2017)<br>94.8 (1990) | 103.2 (2017)<br>201.3 (1990) | 33.1 (2017)<br>136.5 (1990) |
| Maternal mortality (per 100,000)<sup>32</sup> | 178 | 398 | 510 | 814 | 176 |
| Births attended by skilled health personnel (%)<sup>56</sup> | 69% (2014-2019)<br>84% urban (2017-18)<sup>43</sup><br>63% rural (2017-18)<sup>43</sup> | 64% (2014-2019) | 62% (2014-2019) | 43% (2014-2019) | 53% (2014-2019) |

<sup>a</sup> On 01/07/2020, Tanzania was reclassified from ‘low income’ to ‘lower middle income country’ status, by the World Bank, but remains a ‘least developed country’ under the United Nations’ classification system (see https://allafrica.com/stories/202007090579.html)
<sup>b</sup> GNI per capita indicates income status and available resources. LDC inclusion threshold in UN 2018 triennial review was US\$1,025
<sup>c</sup> EVI (0-100) indicates a country’s structural vulnerability to environmental and economic shocks. The EVI threshold for LDC inclusion was 36 in the UN’s 2018 triennial review (the lower the number, the lower the country’s economic vulnerability). Data from: https://www.un.org/development/desa/dpad/least-developed-country-category/ldc-data-retrieval.html (retrieved 31/07/2020)

|  |  |  |  |  |  |
| --- | --- | --- | --- | --- | --- |
| Skilled health professionals density (per 10 000 population)<sup>35</sup> | 14.79 (2015) | 4.38 (2014) | 17.86 (2014) | 18.25 (2009) | 7.38 (2015) |
| Medical doctors (per 10 000 pop)<sup>35</sup> | 9.801 (2018) | 0.14 (2016) | 1.565 (2018) | 3.806 (2018) | 5.809 (2018) |
| Nursing, midwifery personnel (per 10 000 population)<sup>35</sup> | 6.683 (2018) | 5.843 (2017) | 11.656 (2018) | 11.792 (2018) | 4.124 (2018) |
| Community health workers (number) | 95,000 (2020)<sup>60</sup><br>(Lady Health Workers) | 41,000 (2018)<sup>61</sup> | 58,079 (2018)<sup>35</sup> | 116,454 (2018)<sup>35</sup> | 55,136 (2018)<sup>35</sup> |
| Hospital beds (per 10,000 pop)<sup>34</sup> | 6.0 (2015) | 7.0 (2010) | 14.0 (2010) | 5.0 (2004) | 7.7 (2015) |
| Health system financing and service arrangements | Health system devolved (2010) to four largely-autonomous provinces, funded by federal government transfers and development partners, including the World Bank.<sup>62</sup><br>The federal government plays an oversight and coordination role. It manages key vertical programmes, the Lady Health Workers Programme and tertiary hospitals<sup>63</sup>.<br>Pluralist healthcare delivery system comprising public and private (small-scale and commercial for-profit, not-for-profit) and international philanthropies/development stakeholders. Recent policy efforts have been channeled into regulating the private sector and developing public- | Health system (1990s) devolved to semi-autonomous local government authorities and district councils funded by central government transfers, locally-generated revenue and significant funding support from development partners<sup>65</sup>.<br>The national government is largely responsible for policy planning and development, resource mobilization, capacity building, monitoring, evaluation and quality assurance<sup>65</sup>.<br>A district-owned, voluntary Community Health Fund (CHF) (2001), designed for rural and low-income areas, offers members a basic package of health services at primary and hospital levels. Households contribute | Health system devolved (2013) to 47 semi-autonomous counties, funded by central government transfers, locally-generated revenue and donors<sup>67</sup>.<br>Devolution has reduced availability and affordability barriers to care in many counties but less attention has been given to promoting service acceptability and improving care quality<sup>67</sup>.<br>A National Health Insurance Fund (NHIF) offers outpatient and inpatient services, with membership subsidies for the poorest households. The uptake for NHIF stands at 19%<sup>40</sup>.<br>Public-sector services are free at point of use for primary care (dispensaries, | Three-tier system, with primary care decentralized to 774 local government authorities, secondary care managed at state level in 36 states and tertiary care which is the responsibility of the federal government<sup>69</sup>.<br>About 33% (11,395) of health facilities are privately owned and are largely regulated by the State government authorities<sup>70</sup>.<br>National Health Insurance Scheme (NHIS) is a form of social health insurance, operational since 2005, and available to federal formal sector employees (less than 5% enrollment)<sup>71</sup>.<br>Public sector primary care is mostly free at point of use | Decentralized health system without devolution: central decision-making by the Ministry of Health and Family Welfare, operationalized through seven administrative divisions comprising 64 Zila (districts) and 492 Upazila (sub-districts), each with one Upazila Health Complex<sup>72</sup>.<br>Pluralist and largely unregulated healthcare system, comprising public, private (individual/small-scale-for-profit, commercial, not-for-profit) and international philanthropies/development stakeholders. Three-tiered public primary healthcare (PHC) system delivers basic preventive and curative services at Community Clinics (village level), Union Health and Family Welfare Centres at |
|  | private partnerships for health<sup>60</sup>.<br>Health insurance is not mandatory and voluntary health insurance is negligible (0.2% of national health expenditure). Khyber Pukhtoonkhwa Province has recently introduced micro health insurance through the Sehat Sahulat Program.<sup>41</sup><br>Public health care is free at point of use for all primary, secondary and tertiary care (except for certain specialized tests that may incur a nominal fee).<br>Pakistan is 1 of 57 countries with a critical health workforce deficiency, according to WHO<sup>64</sup>. | annual fees to the CHF, with match-funding from the central government<sup>65</sup> (4% coverage).<sup>39</sup><br>The National Health Insurance Fund (NHIF) is mandatory for government employees and open to other formal sector employees and those who can afford to pay<sup>65</sup> (17% coverage)<sup>39</sup>.<br>Primary healthcare (PHC) is provided through small dispensaries (basic preventive and curative services, approx 15-20 providers) and bigger health centres (inpatient and higher level services, staffed by more and better-qualified healthcare providers (approx.39-52). Referrals are to the district hospital.<sup>66</sup><br>Since 1993, user fees have been charged for public PHC but children<5 and those aged 60+ are exempted. | health centres) and all maternal care<sup>68</sup>. |  | the Union level (collection of villages) and Upazila Health Complexes (sub-district level, provide limited inpatient services). District hospitals provide secondary level care, with tertiary hospitals in large urban centres<sup>73, 74</sup><br>There is no mandatory health insurance. Voluntary schemes (public and private) are negligible<sup>74</sup>.<br>Primary public care: is free at point of use for outpatient services (beyond a nominal patient registration fee). Public sector hospital fees are minimal. |
| Universal Health Coverage index of essential service coverage<sup>e34</sup> | 0.45 (2017)<br>0.42 (2015) | 0.43 (2017)<br>0.41 (2015) | 0.55 (2017)<br>0.54 (2015) | 0.42 (2017)<br>0.42 (2015) | 0.48 (2017)<br>0.46 (2015) |
| Out-of-pocket (OOPS) as % of Current Health Expenditure<sup>37</sup> | 60% (2017)<br>64% (2012) | 24% (2017)<br>25% (2012) | 24 % (2017)<br>32% (2012) | 77% (2017)<br>73% (2012) | 74% (2017)<br>67% (2012) |
| Top 10 causes of most death and disability combined (2017)<sup>32</sup> | Neonatal disorders, ischemic heart disease, lower respiratory infection, diarrhoeal diseases, road injuries, stroke, congenital defects, TB, diabetes, cirrhosis | Neonatal disorders, lower respiratory tract, HIV/AIDS, congenital defects, malaria, TB, diarrhoeal disease, protein-energy malnutrition, ischemic heart disease, dietary iron deficiency | HIV/AIDS, neonatal disorders, diarrhoeal diseases, lower respiratory infection, congenital defects, TB, stroke, ischemic heart disease, meningitis, cirrhosis | Neonatal disorders, lower respiratory tract, malaria, diarrhoeal diseases, HIV/AIDS, meningitis, congenital defects, TB, protein-energy malnutrition, dietary iron deficiency | Neonatal disorders, stroke, ischemic heart disease, lower respiratory infection, COPD, low back pain, headache disorders, diabetes, diarrhoeal diseases, congenital defects |
| Top 5 risk factors driving the most death and disability (2017)<sup>32</sup> | Malnutrition, dietary risks, high blood pressure, tobacco, air pollution | Malnutrition, unsafe sex, air pollution, WASH, high blood pressure | Malnutrition, unsafe sex, WASH, alcohol use, air pollution | Malnutrition, WASH, air pollution, unsafe sex, high blood pressure | Malnutrition, dietary risks, air pollution, high blood pressure, tobacco |
| Electricity | 2017-2018<sup>43</sup><br>National: 93%<br>Urban: 99%<br>Rural: 89% | 2016<sup>42</sup><br>National: 32.8 %<br>Urban: 65.3%<br>Rural: 16.9% | 2014<sup>75</sup><br>National: 36%<br>Urban: 68%<br>Rural: 13% | 2018<sup>76</sup><br>National: 59.4%<br>Urban: 82.7%<br>Rural: 38.9% | 2017-2018<sup>77</sup><br>National: 90.9%<br>Urban: 96.5%<br>Rural: 88.7% |
| Mobile phone subscriptions in millions (% pop)<sup>44</sup> | 164.7 (75%) (2020)<br>+6.2% (since 2019) | 44.13 (75%) (2020)<br>+1.6% (since 2019) | 52.06 (98%) (2020)<br>+8.7% (since 2019) | 169.2 (83%) (2020)<br>+7.7% (since 2019)<br>(88% in 2018 DHS)<sup>76</sup> | 163 (99%) (2020)<br>+4.5% (since 2019)<br>(94.4% in 2017-18 DHS)<sup>77</sup> |
| Mobile phone ownership (gender/ place of residence) | 2017-2018<sup>43</sup><br>Men: 93%<br>Women: 39%<br>Sindh rural: 5.7% of women (15-49) own a mobile phone | 2015-2016<sup>45</sup><br>Men: 82% (urban), 74% (rural)<br>Women: 62% (urban), 40% (rural) | 2014<sup>75</sup><br>Urban: 94%, Rural: 80% (no gender breakdown available in DHS) | 2018<sup>76</sup><br>Urban households: 94.5%<br>Rural households: 82.1%<br>Men: 81% (age 15-49)<br>Women: 55% (age 15-49) | 2017-2018<sup>77</sup><br>Urban: 96.6%, Rural: 93.6%<br>Men (unmarried, 15-19): 65% (urban), 63.6% (rural)<br>Women (unmarried, 15-19): 39% (urban), 30% (rural)<br>Women (married, 15-49): 69.6% (urban), 55.6% (rural) |
| Internet penetration (users in millions)<sup>44</sup> | 76.38 (35%) (2020) | 14.72 (23%) (2020) | 22.86 (43%) (2020) | 85.49 (42%) (2020) | 66.4 (41%) (2020) |
| Internet usage in the past 12 months (gender/place of residence) | 2017-2018<sup>43</sup><br>National: 28.4% men, 12.0% women<br>Sindh rural: 9.9% men, 1.5% women<br>4.9% rural households have internet connection | 2015-2016<sup>45</sup><br>National: 19% men, 8% women<br>Rural: 9% men, 2 % women | No information on internet usage in 2014 DHS | 2018<sup>76</sup><br>Men: 55% (urban)<br>25% (rural)<br>Women: 31% (urban)<br>6% (rural) | No information on internet usage in 2017-2018 DHS |
| Digital and eHealth-related policies | eHealth is embedded in the national *Digital Pakistan* (2018) policy<sup>47</sup>. | *Tanzania National eHealth Strategy June, 2013 – July, 2018 (2013)* focuses on improving broadband services<sup>51</sup>;<br>*National Five Year Development Plan 2016/17 – 2020/21 (2016)*: includes mHealth apps<sup>78</sup>;<br>*Digital Health Investment Roadmap (2017-2023)* concentrates on data use for improving system performance<sup>48</sup>. | eHealth/mHealth are embedded in national policies. Specific policies: *Standards and Guidelines for mHealth Systems* (2017)<sup>52</sup> and the promotion of service digitization and technology adoption including mHealth, for *Transforming Lives* (2018-2022)<sup>53</sup>. There is a dedicated *eHealth Development Unit* but this is not joined up to the Ministry of ICT. | No specific national mHealth/ eHealth policy but the *National Health ICT Strategic Framework 2015 - 2020* visualises that “By 2020, health ICT will help enable and deliver universal health coverage”<sup>46</sup>. | eHealth/mHealth are embedded in national policies. Specific policies: *Draft Guidelines for eHealth Standards*<sup>54</sup> and an interoperability framework, to reduce duplication of effort and facilitate linkages among mHealth initiatives. *Digital Bangladesh Vision 2021*: healthcare is a priority<sup>49</sup>. The *Directorate General of Health Services* seeks to improve access and availability of digital health services. |
| | | East African Community: *Health Sector Investment Priority Framework* (2018 - 2028) - sub priority 9.3: Investment in Digital Health Technology for better research for health, health services delivery and health outcomes has a budget of US\$ 21,175,000 for estimated cost of US\$3,175, 699, 500)<sup>55</sup> | | | |

In the last two decades, each country has made progress in increasing life expectancy, improving maternal and child health outcomes and reducing malnutrition. However, malnutrition remains the biggest risk factor for death and disability in all^32^, and maternal mortality rates in Nigeria, Kenya and Tanzania are substantially higher than the WHO/Sustainable Development Goal of 140 per 100,000 live births^33^. All five countries are working towards universal health coverage^34^, but face challenges of weak health systems, shortages in skilled health workers^35^ and a growing burden of non-communicable disease, alongside an already-high burden of communicable disease^32^. There are differences in health system financing and service arrangements but all are pluralist, involving a complex mix of public and private sector services, spanning individual/small for-profit, commercial and not-for-profit stakeholders. International development partners are key funding stakeholders, particularly in Pakistan, Bangladesh and Tanzania, including for digital health^36^. Public sector primary care is free at point of use in Pakistan, Kenya, Nigeria (mostly) and Bangladesh, while user fees (with some exemptions) apply in Tanzania. Out-of-pocket healthcare expenditure is high in Bangladesh (74%), Nigeria (77%) and Pakistan (60%), lower in Tanzania (24%) and Kenya (24%)^37^. In Kenya, almost 7% of the population is pushed into poverty, annually, as a result of direct payments for healthcare and associated transport costs.^38^ Health insurance is negligible in Bangladesh and Nigeria, while various social and voluntary health insurance schemes have some traction in Tanzania^39^ and Kenya^40^, including in rural areas. A micro health insurance system, to support ‘under-privileged citizens’ to access needed healthcare, has recently been introduced in Pakistan’s Khyber Pukhtoonkhwa Province^41^. Additional country contrasts include infrastructure and access to electricity (e.g., in Tanzania, access is 32.8% national/16.9% rural42 compared to 93% national/89% rural in Pakistan^43^), and diverse socio-political history and culture.

In keeping with global trends^44^, mobile phone subscriptions are high in all five countries, ranging from 75% of the population in Tanzania and Pakistan, to almost 100% in Kenya and Bangladesh (Table 1). Internet penetration is lower in all - between 23% in Tanzania and just over 40% in Nigeria, Bangladesh and Kenya^44^. In each country, there are gender and location differences in mobile phone ownership and internet usage, indicating less independent access for women and rural residents. For example, 93% men and only 39% women own mobile phones in Pakistan^43^. In urban Tanzania, 82% men, 62% women own mobile phones compared to 74% men, 40% women in rural areas^45^.

In all five countries, technology-enabled healthcare delivery is embedded in national policies, including on Information and Communication Technology (ICT) (Nigeria^46^) and digital futures (Pakistan^47^, Tanzania^36,48^Bangladesh^49,50^). Specific electronic/eHealth and mobile/mHealth policies are in place in Tanzania^51^, Kenya^52, 53^ and Bangladesh^54^ (Table 1). Kenya has developed standards and guidelines for mHealth systems (2017)^52^, and in Nigeria, there are efforts to incorporate and regulate ICT through existing health policies, including those that govern face-to-face consultation (e.g., professionalism, confidentiality). Regionally, as members of the East African Community, Kenya and Tanzania are guided by the Health Sector Investment Priority Framework (2018 - 2028), which promotes investment in digital health technology^55^.

### Study setting: Communities with minimal access to healthcare services

Within the five countries, we undertook a scoping study of mConsulting: in remote rural areas in Pakistan and Tanzania, and urban slums in Bangladesh, Kenya and Nigeria. These sites were, purposively, selected as low resource communities with minimal access to healthcare services, located in rural-urban contrast. The five urban sites form part of the NIHR Global Health Research Unit on Improving Health in Slums study^79^. Their inclusion gave us access to secondary data from household and adult surveys (conducted in 2018-19), which asked questions on mobile phone access, internet access, and digital health-seeking behaviour^80, 81^. Table 2 provides a description of each site, including contextualising information for the urban sites from the NIHR Global Health Research Unit on Improving Health in Slums^81, 82^.

**Table 2.**
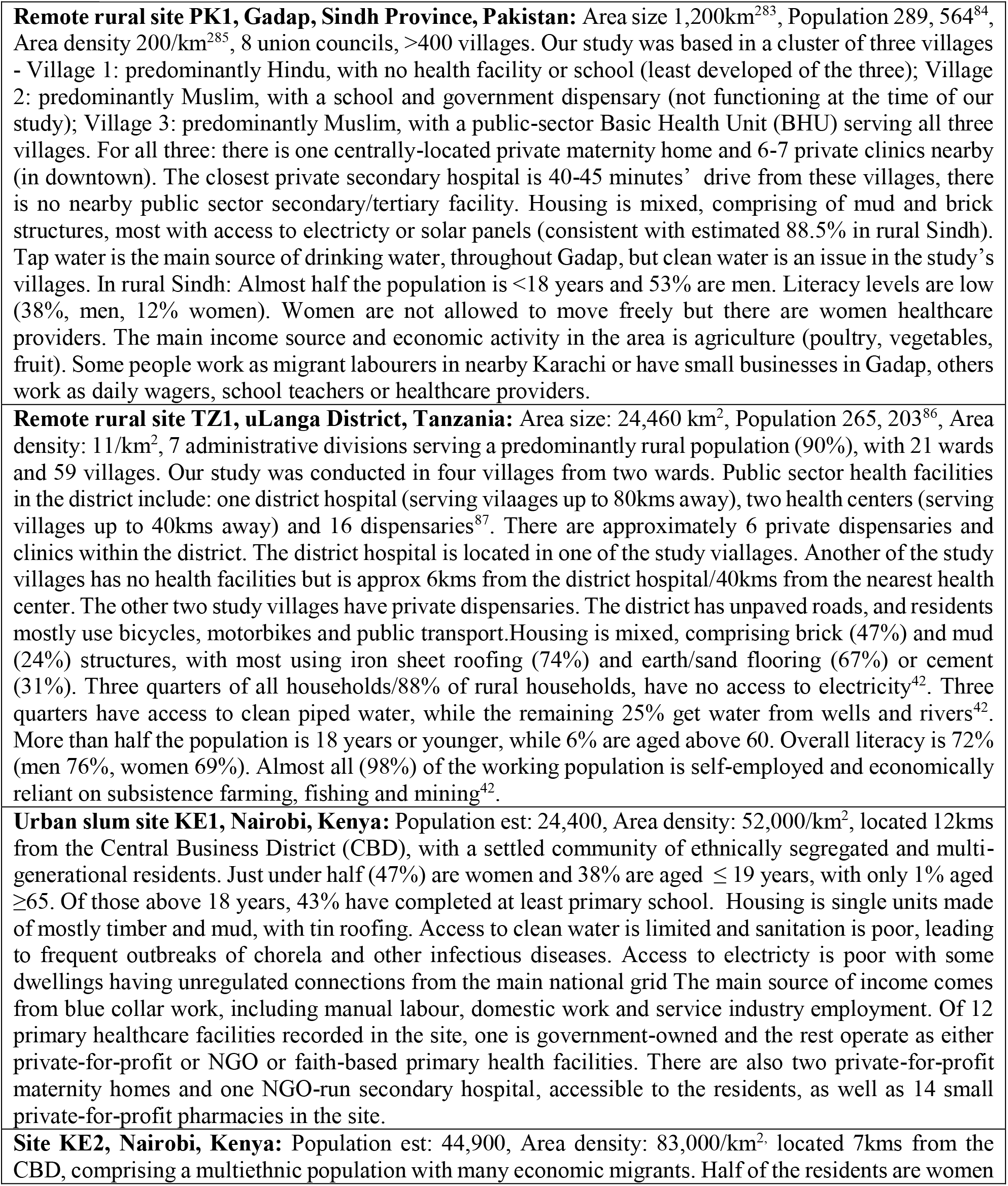
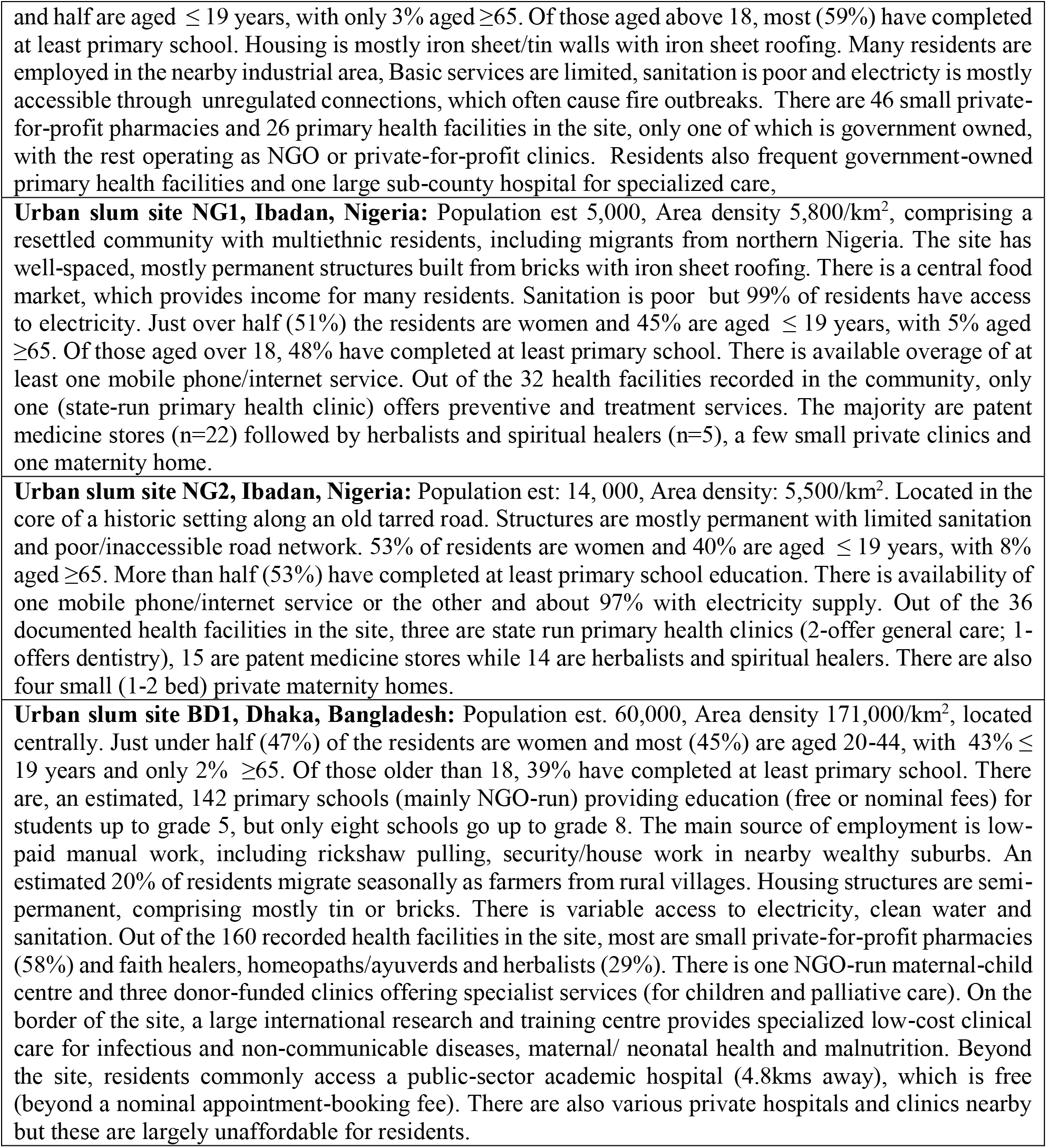
Study sites: Low-resource communities with minimal access to healthcare.

#### Remote rural site PK1, Gadap, Sindh Province, Pakistan

Area size 1,200km^283^, Population 289, 564^84^, Area density 200/km^285^, 8 union councils, >400 villages. Our study was based in a cluster of three villages - Village 1: predominantly Hindu, with no health facility or school (least developed of the three); Village 2: predominantly Muslim, with a school and government dispensary (not functioning at the time of our study); Village 3: predominantly Muslim, with a public-sector Basic Health Unit (BHU) serving all three villages. For all three: there is one centrally-located private maternity home and 6-7 private clinics nearby (in downtown). The closest private secondary hospital is 40-45 minutes’ drive from these villages, there is no nearby public sector secondary/tertiary facility. Housing is mixed, comprising of mud and brick structures, most with access to electricty or solar panels (consistent with estimated 88.5% in rural Sindh). Tap water is the main source of drinking water, throughout Gadap, but clean water is an issue in the study’s villages. In rural Sindh: Almost half the population is <18 years and 53% are men. Literacy levels are low (38%, men, 12% women). Women are not allowed to move freely but there are women healthcare providers. The main income source and economic activity in the area is agriculture (poultry, vegetables, fruit). Some people work as migrant labourers in nearby Karachi or have small businesses in Gadap, others work as daily wagers, school teachers or healthcare providers.

#### Remote rural site TZ1, uLanga District, Tanzania

Area size: 24,460 km^2^, Population 265, 203^86^, Area density: 11/km^2^, 7 administrative divisions serving a predominantly rural population (90%), with 21 wards and 59 villages. Our study was conducted in four villages from two wards. Public sector health facilities in the district include: one district hospital (serving vilaages up to 80kms away), two health centers (serving villages up to 40kms away) and 16 dispensaries^87^. There are approximately 6 private dispensaries and clinics within the district. The district hospital is located in one of the study viallages. Another of the study villages has no health facilities but is approx 6kms from the district hospital/40kms from the nearest health center. The other two study villages have private dispensaries. The district has unpaved roads, and residents mostly use bicycles, motorbikes and public transport.Housing is mixed, comprising brick (47%) and mud (24%) structures, with most using iron sheet roofing (74%) and earth/sand flooring (67%) or cement (31%). Three quarters of all households/88% of rural households, have no access to electricity^42^. Three quarters have access to clean piped water, while the remaining 25% get water from wells and rivers^42^. More than half the population is 18 years or younger, while 6% are aged above 60. Overall literacy is 72% (men 76%, women 69%). Almost all (98%) of the working population is self-employed and economically reliant on subsistence farming, fishing and mining^42^.

#### Urban slum site KE1, Nairobi, Kenya

Population est: 24,400, Area density: 52,000/km^2^, located 12kms from the Central Business District (CBD), with a settled community of ethnically segregated and multi-generational residents. Just under half (47%) are women and 38% are aged ≤ 19 years, with only 1% aged ≥65. Of those above 18 years, 43% have completed at least primary school. Housing is single units made of mostly timber and mud, with tin roofing. Access to clean water is limited and sanitation is poor, leading to frequent outbreaks of chorela and other infectious diseases. Access to electricty is poor with some dwellings having unregulated connections from the main national grid The main source of income comes from blue collar work, including manual labour, domestic work and service industry employment. Of 12 primary healthcare facilities recorded in the site, one is government-owned and the rest operate as either private-for-profit or NGO or faith-based primary health facilities. There are also two private-for-profit maternity homes and one NGO-run secondary hospital, accessible to the residents, as well as 14 small private-for-profit pharmacies in the site.

#### Site KE2, Nairobi, Kenya

Population est: 44,900, Area density: 83,000/km^2,^ located 7kms from the CBD, comprising a multiethnic population with many economic migrants. Half of the residents are women and half are aged ≤ 19 years, with only 3% aged ≥65. Of those aged above 18, most (59%) have completed at least primary school. Housing is mostly iron sheet/tin walls with iron sheet roofing. Many residents are employed in the nearby industrial area, Basic services are limited, sanitation is poor and electricty is mostly accessible through unregulated connections, which often cause fire outbreaks. There are 46 small private-for-profit pharmacies and 26 primary health facilities in the site, only one of which is government owned, with the rest operating as NGO or private-for-profit clinics. Residents also frequent government-owned primary health facilities and one large sub-county hospital for specialized care,

#### Urban slum site NG1, Ibadan, Nigeria

Population est 5,000, Area density 5,800/km^2^, comprising a resettled community with multiethnic residents, including migrants from northern Nigeria. The site has well-spaced, mostly permanent structures built from bricks with iron sheet roofing. There is a central food market, which provides income for many residents. Sanitation is poor but 99% of residents have access to electricity. Just over half (51%) the residents are women and 45% are aged ≤ 19 years, with 5% aged ≥65. Of those aged over 18, 48% have completed at least primary school. There is available overage of at least one mobile phone/internet service. Out of the 32 health facilities recorded in the community, only one (state-run primary health clinic) offers preventive and treatment services. The majority are patent medicine stores (n=22) followed by herbalists and spiritual healers (n=5), a few small private clinics and one maternity home.

#### Urban slum site NG2, Ibadan, Nigeria

Population est: 14, 000, Area density: 5,500/km^2^. Located in the core of a historic setting along an old tarred road. Structures are mostly permanent with limited sanitation and poor/inaccessible road network. 53% of residents are women and 40% are aged ≤ 19 years, with 8% aged ≥65. More than half (53%) have completed at least primary school education. There is availability of one mobile phone/internet service or the other and about 97% with electricity supply. Out of the 36 documented health facilities in the site, three are state run primary health clinics (2-offer general care; 1-offers dentistry), 15 are patent medicine stores while 14 are herbalists and spiritual healers. There are also four small (1-2 bed) private maternity homes.

#### Urban slum site BD1, Dhaka, Bangladesh

Population est. 60,000, Area density 171,000/km^2^, located centrally. Just under half (47%) of the residents are women and most (45%) are aged 20-44, with 43% ≤ 19 years and only 2% ≥65. Of those older than 18, 39% have completed at least primary school. There are, an estimated, 142 primary schools (mainly NGO-run) providing education (free or nominal fees) for students up to grade 5, but only eight schools go up to grade 8. The main source of employment is low-paid manual work, including rickshaw pulling, security/house work in nearby wealthy suburbs. An estimated 20% of residents migrate seasonally as farmers from rural villages. Housing structures are semi-permanent, comprising mostly tin or bricks. There is variable access to electricity, clean water and sanitation. Out of the 160 recorded health facilities in the site, most are small private-for-profit pharmacies (58%) and faith healers, homeopaths/ayuverds and herbalists (29%). There is one NGO-run maternal-child centre and three donor-funded clinics offering specialist services (for children and palliative care). On the border of the site, a large international research and training centre provides specialized low-cost clinical care for infectious and non-communicable diseases, maternal/ neonatal health and malnutrition. Beyond the site, residents commonly access a public-sector academic hospital (4.8kms away), which is free (beyond a nominal appointment-booking fee). There are also various private hospitals and clinics nearby but these are largely unaffordable for residents.

### Study design

Set within the seven study sites described in Table 2, our scoping study involved: 1) policy and document review; 2) secondary quantitative analysis of data from household and adult surveys, undertaken by the NIHR Global Health Research Unit on Improving Health in Slums^79^ in the five urban study sites; followed by 3) qualitative interviews and workshops with key stakeholders in all study sites (urban slums and remote rural areas). For the urban sites, we were able to mix our methods in explanatory design,^88^ first identifying the extent of mConsulting through the surveys and then exploring with stakeholders how representative the survey findings are.^89^ For all study sites, we designed our qualitative engagements to include multiple perspectives from within mConsulting systems; 4) our approach was guided by project advisory groups, comprising community representatives, local health workers and mConsulting providers, in each country; 5) in our interpretation, we integrated our findings both within this scoping study and with our review of current evidence, in an effort to capture a wider ‘picture of a system’ - a complex, adaptive mConsulting system - informed by multiple perspectives^89^.

Ethical clearance and approvals were obtained from all relevant bodies in each partner institution and study site. All participants provided informed consent.

### Secondary data collection

#### Household and adult survey data (NIHR Global Health Research Unit on Improving Health in Slums)^80^

Between 2018 and 2019, household and adult surveys were conducted by the Unit in the five urban slum sites, using a geospatially-referenced study design and survey methods that have been described elsewhere^80, 81^. Administered by fieldworkers trained in the ethics and techniques of survey-based data collection, in the language preferred by the respondent, household surveys were used to collect demographic and socioeconomic data, while individual surveys (administered in each participating household to a randomly selected household resident aged over 18), collected health-related information. For the purposes of our study, we used data collected about household access to mobile phones, internet and airtime; and adult digital health-seeking behaviour (see Appendix 2).

### Primary data collection

#### Selection and recruitment of participants

We purposefully selected participants for their role in the mConsulting system: policy-makers and digital health experts, telecommunication providers, mConsulting providers, health workers and community members. We identified mConsulting service providers and users through internet searches, our organisational networks, site contacts and by word-of-mouth. Health care workers were drawn from cadres active in local care provision, including clinical officers, doctors, nurses, pharmacists and community health workers.

We selected residents for diversity of age, gender and religion, choosing different times of the day across the working week/weekends, and different part of each site to reach people ‘at home’. In the urban study sites, we used the findings from our secondary analysis of the surveys to contact trace community members (who had used their mobile phones to receive health information/advice and who had agreed to participate in follow-on studies). These participants were invited to participate in mini-interviews and community workshops. Health workers and decision-makers were identified through previous engagements, site contacts and, in the urban sites, from previous mapping of healthcare facilities undertaken as part of the NIHR Global Health Research Unit on Improving Health in Slums^79^.

We reviewed policies about mConsulting and interviewed policy and digital experts. We held community workshops and interviews to ask community leaders, local healthcare workers, pharmacists, shop and drug vendors, and other community members about mConsulting services, exploring what is available, used and why? We interviewed mConsulting providers about their purpose, history, size and coverage, operating systems and costs. With all participants, we explored their perceptions of the impact of mConsulting on users and the health system and sought their ideas about whether mConsulting is an option to strengthen access to health care.

Towards the end of the study, we brought together community members, health workers, mConsulting service providers and decision-makers for consensus-building workshops to discuss our findings and develop ideas for health policy and for future research.

Interviews and workshops were carried out at venues convenient and accessible to participants, in their preferred language. They were conducted by researchers trained in the methods and ethics of qualitative engagements, including taking of consent. Semi-structured interview guides were piloted and refined following feedback.

Regular debriefing sessions were held with researchers to identify issues for further exploration and to manage any unanticipated problems.

For community-level mini-interviews, informed verbal consent was sought from participants and noted in field notes. Field notes included the participants’ role in the community and any mConsulting services mentioned. These were typed up and expanded by the researcher in English, as soon as possible, after each interview. For the semi-structured interviews with healthcare providers, key informants and policy/decision-makers, informed written consent was sought, including to audio-record the interview.

All identifiers were removed from transcripts and quality checked by research team members. Data were encrypted and stored on a secure server at the University of Warwick for analysis.

### Data analysis

#### Secondary analysis of household and adult survey data (NIHR Global Health Research Unit on Improving Health in Slums)^80^

Researchers in each country-team (MA, NC, PK) tabulated data from the relevant sections of the household and adult surveys (mobile phone, internet, airtime access, use of technology for health-care seeking) (Appendix 2). For each site, the total sample for that site was tabulated against the total number of respondents for that particular question per site.

### Qualitative analysis of interviews and community workshops

Interviews and field notes were transcribed and translated into English where necessary. Transcripts were reviewed by team members (BC, MA, PK, RA) against audio recordings to ensure accuracy of translations and consistency. These were analysed thematically^90^, guided by our understanding of access as a dynamic interchange between mConsulting users and providers across a digital communication platform.^9, 10^ Researchers in each country-team (BC, OF, MA, PB, PK, NR, NC) coded the transcripts along key access dimensions of acceptability, availability and affordability, while allowing for emergent themes. In consultation with the wider team, codes were reviewed, then developed into initial themes and refined through further coding. Themes were compared across countries and according to participant-type.

### Patient and public involvement

At the beginning of our community-based research, we consulted with community leaders in each site. Community members are part of Project Advisory Groups (PAGs) in each country team. The PAGs have advised us on our research approach, process and plans, including dissemination of results. Community members were recruited to fieldwork teams in Nigeria, Kenya and Bangladesh. Members of the community, including patients, were included as study participants.

## Results

Between August 2019 and March 2020, we collected primary data from 419 participants: We carried out interviews (approx. 30-60 minutes each) with mHealth policy experts, telecommunication providers and mConsulting providers (50), community decision makers (9), local health workers (34) and community members (144). We held 12 community workshops (approx. 2-3 hours each), attended by 121 residents and local health workers and, in Nigeria and the two Kenyan sites, we held three half-day consensus workshops reaching 61 decision makers, health workers and residents. During the study period, eight project advisory meetings were held across the sites. COVID-19 disrupted some of our planned project activities. In Bangladesh, we postponed interviews with policy-makers, digital experts and mConsulting service providers; in Pakistan, Tanzania and Bangladesh, we cancelled consensus workshops. In all sites, we used research briefings and continued engagement with project advisors to deliver feedback and disseminate our findings. Table 3 provides a breakdown of the activities and participation in each country. Additionally, we tabulated data from household and adult survey datasets collected in each of the urban slum study sites (total n=5,219 households, n=5,186 adults drawn from the households surveyed) as part of the NIHR Global Health Research Unit on Improving Health in Slums (Table 4) ^79-81^.

**Table 3.**
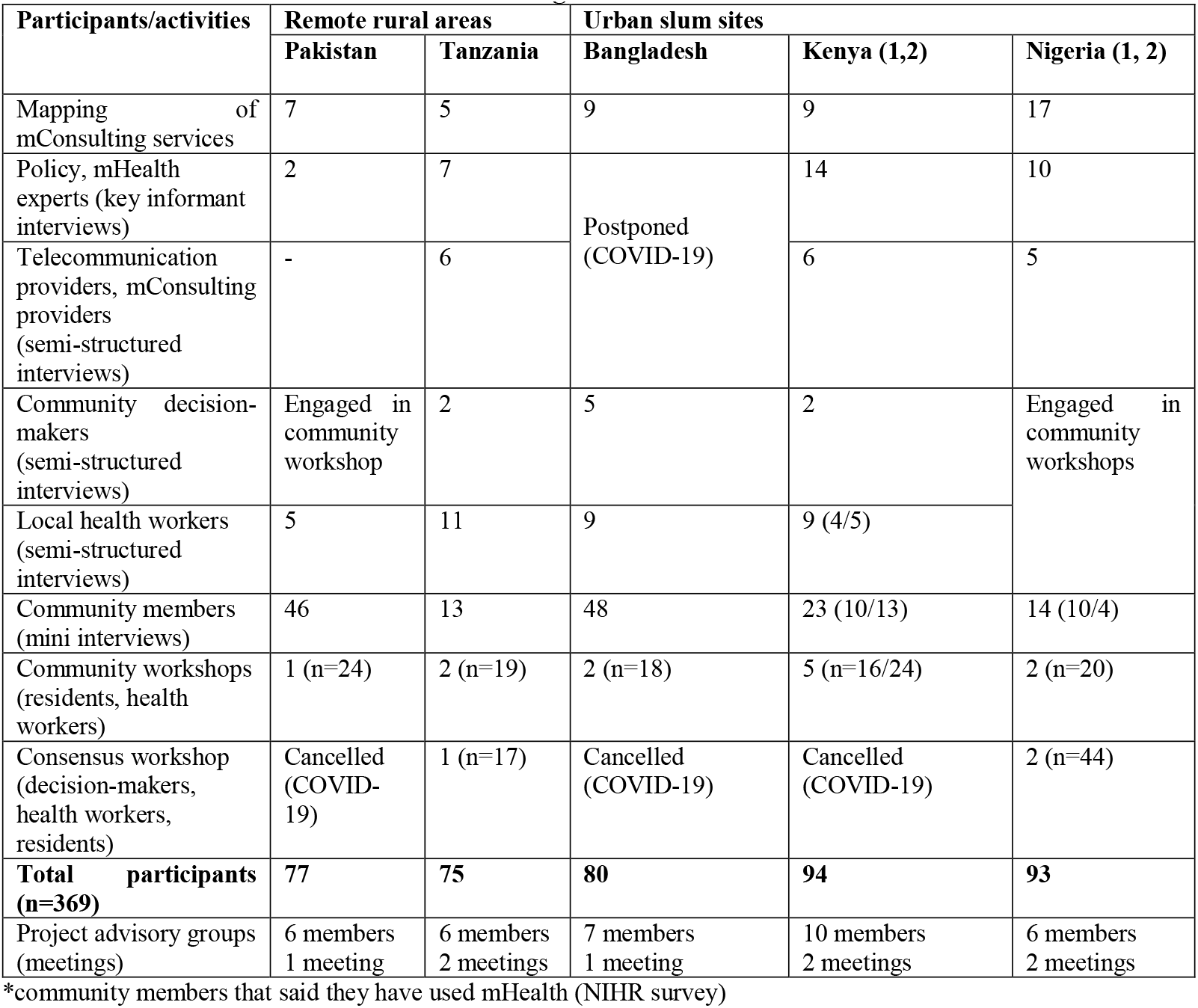
Activities undertaken between August 2019 and March 2020.

| Participants/activities | Remote rural areas |  | Urban slum sites |  |  |
| --- | --- | --- | --- | --- | --- |
|  | Pakistan | Tanzania | Bangladesh | Kenya (1,2) | Nigeria (1, 2) |
| Mapping of mConsulting services | 7 | 5 | 9 | 9 | 17 |
| Policy, mHealth experts (key informant interviews) | 2 | 7 | Postponed (COVID-19) | 14 | 10 |
| Telecommunication providers, mConsulting providers (semi-structured interviews) | - | 6 |  | 6 | 5 |
| Community decision-makers (semi-structured interviews) | Engaged in community workshop | 2 | 5 | 2 | Engaged in community workshops |
| Local health workers (semi-structured interviews) | 5 | 11 | 9 | 9 (4/5) |  |
| Community members (mini interviews) | 46 | 13 | 48 | 23 (10/13) | 14 (10/4) |
| Community workshops (residents, health workers) | 1 (n=24) | 2 (n=19) | 2 (n=18) | 5 (n=16/24) | 2 (n=20) |
| Consensus workshop (decision-makers, health workers, residents) | Cancelled (COVID-19) | 1 (n=17) | Cancelled (COVID-19) | Cancelled (COVID-19) | 2 (n=44) |
| **Total participants (n=369)** | **77** | **75** | **80** | **94** | **93** |
| Project advisory groups (meetings) | 6 members<br>1 meeting | 6 members<br>2 meetings | 7 members<br>1 meeting | 10 members<br>2 meetings | 6 members<br>2 meetings |

\*community members that said they have used mHealth (NIHR survey)

**Table 4.** Household access to mobile phones, the internet and individual use of mConsulting in the urban study sites.

|  | Nigeria |  | Kenya |  | Bangladesh |
| --- | --- | --- | --- | --- | --- |
|  | Site 1 | Site 2 | Site 1 | Site 2 |  |
| Number of households (total n=5,219) | 845 | 1,286 | 1,018 | 1,080 | 990 |
| Household has access to a mobile phone (Yes) | 85% | 87% | 87% | 95% | 97% |
| Available airtime: Every day | 19% | 15% | 16% | 29% | 78% |
| Access to data/WiFi: Never | 77% | 66% | 37% | 48% | 56% |
| Number of adults (total n = 5,186) | 834 | 1,264 | 1018 | 1080 | 990 |
| In the last 12 months have you used or attempted to use your mobile phone or other digital communication device (e.g.: laptop, | 0.7%<br>(n=6) | 1.4%<br>(n=18) | 2.5%<br>(n=25) | 3.4%<br>(n=37) | 0.2%<br>(n=2) |
| tablet) to access health information, advice or care for yourself, where information about your health was received or given? (n=Yes adult respondents) (Total n=88) |  |  |  |  |  |
| Qualitative follow-up - those who said Yes | - | 5 | 17 | 19 | - |

Source: Surveys (household and adult) NIHR Global Health Research Unit on Improving Health in Slums

### Access to mobile devices and connectivity

Consistent with the rapid rise in mobile communication technology in LMICs over the last decade, 75-99% of the population in the study countries has mobile phone subscriptions, as shown in Table 1^44^. From our analysis of the household data in the urban sites, most households (85% and more) reported access to a mobile phone (almost 100% in Kenya 2 and Bangladesh) (Table 4).

In Bangladesh, 4 in 5 households had access to airtime every day, compared to fewer than 1 in 5 in Nigeria 2 and Kenya 1 (Table 4). Through our interviews and community workshops, participants in the rural sites confirmed that their households owned or could borrow a basic mobile phone, although fewer had access to a smartphone. In urban Kenya, participants told us that more women owned smartphones. In both rural sites, we were told that more men than women owned phones, which is consistent with data collected by the Demographic and Health Surveys, in each country^43,45^. In Tanzania, lack of reliable electricity was identified as a barrier to device use.

Unlike mobile phone access, far fewer households, nationally^44^ (see Table 1) and in the study sites, had regular, if any, access to data/WiFi and the internet, with the majority of urban households reporting no access at all, particularly in Nigeria (Table 4). Limited network coverage was raised as a key issue in both rural sites, although residents explained they could usually walk to connection hotspots in their villages.

### Use of mobile phones ‘where information about your health was received or given’ (urban slum sites)

Most households, surveyed in the urban slum sites, had access to a mobile phone. However, only a small number of adult respondents (total n=88) reported that they had used their phone or another digital device to access and receive health information, advice or care in the last 12 months (Table 4). Most of these respondents were living in households in the two Kenyan sites. On qualitative follow-up, survey respondents who said ‘yes’ in Nigeria and Kenya explained that they used their devices to read/post health-related questions on social media groups, and/or contact a known healthcare provider or a medically-trained family member to discuss symptoms and get drug advice/prescriptions:

> *She is on a Facebook group* […] *Members of the group asks questions on health and are directed on how to treat themselves. She has never asked questions on the group but she reads from others. She also has a doctor who she chats with on whatsapp about her health* (*field notes from community workshop, Nigeria*).

> *Calls a nurse* [*name*] *whenever he notices any symptoms. He tells her his symptoms and then she tells him what drug to buy. Sometimes, she comes over to treat him. Respondent can’t remember the hospital where she works* (*field notes from community workshop, Nigeria*).

### Availability of mConsulting services

In presenting our findings, we distinguish between two types of mConsulting services:

a. Those delivered through nationally/regionally-available provider platforms run by commercial companies, government agencies or NGOs, using written communication (text messaging, app-based information, web chats) audio and/or video channels. Consultants include real people and algorithm-driven computers; and
b. mConsulting undertaken by local healthcare workers using their phones to speak to community members using their phones. Health workers include pharmacists, community health workers, nurses, clinical officers and doctors.

#### (a) Nationally/regionally-available mConsulting platforms

We identified between 5 and 17 services operating through provider platforms in each country (Table 5). Many were targeted at specific health conditions or groups: reproductive, maternal-child health, HIV/TB, youth, elderly, and, in Bangladesh and Tanzania, rural areas. Others were available for the general public or unspecified health issues. Some kept patient records. Some offered referrals, follow up services and drug prescriptions. In all but Tanzania, the most common communication channel was text messaging and written communication using web chats or managed through apps, for example, the mDaktari (Kenya) app https://connectmed.co.ke/ provides daily coaching for patients with chronic conditions (hypertension, diabetes) and facilitates online bookings, consultations, prescriptions, referrals and record-keeping through each patient’s account; JamboMama (Tanzania) https://smartaccesstohealthforall.org/jambomama-2/ provides personalised information, advice and monitoring for pregnant women, connecting users (through their encrypted data) to health workers who are able to monitor patient progress and alert them of any ‘predefined unusual, out of safety range data’. Written communication was followed by audio calls (most common at 70% in Tanzania). For a few services, in each country, video was an added option.

**Table 5.** Provider platform services identified in Kenya, Nigeria, Tanzania, Pakistan and Bangladesh (to end-March 2020)

| Services identified<sup>1</sup> | Digital channel used (may be >1 per service) |  |  |  | Consult fee charged | Targeted population /condition<sup>^</sup> | Refer | Keep record | Prescribe drugs | Follow up |
| --- | --- | --- | --- | --- | --- | --- | --- | --- | --- | --- |
|  | App | WC | Audio | Video |  |  |  |  |  |  |
| **Tanzania (5)** | 1 | 1 | 5 |  | 2 fee, 3 free | 1RH, 1MH, 1TB, 1HIV, 2All | 5 | 5 | 4 | DK |
| **Pakistan (7)** | 1 | 4 | 2 | - | 6 fee, 1 free | 2 MH, 5 All | 2 | 2 | 4 | DK |
| **Bangladesh (17)** | 4 | 11 | 10 | 2 | 14 fee, 2 free, 1 DK | 2MH, 3rural, 8 phone company subscriber-specific, 4All | 12 | 5 | 12 |  |
| **Kenya (9)** | 6 | 7 | 7 | 3 | 6 fee, 3 free | 1RH, 5All | 6 | 2 | 1 | DK |
| **Nigeria (17)** | 12 | 13 | 10 | 12 | 14 fee, 3 free | 1MH, 1>age 16, 1elderly 14All | 8 | 9 | 6 | 5 |

<sup>1</sup>WC: web chat (text), RH: reproductive health, MH: maternal health, All: general population, DK: don't know
**Tanzania** (n=5, 2 fee, 5 free) JamboMama, AfyaCall, Tambua TB, Daktari Mkononi, Afya Helpline
**Pakistan** (n=7), 6 fee, 1 free: MDConsults/MyMDConsults (Tech4Life), Augmentcare: Marham, MyZindagi(ORG), Sehatyab, DoctHERS. Sehat Kahani, SUKH initiative
**Bangladesh** (n=17 services delivered by 7 providers, 14 fee, 2 free, 1 DK): (DGHS) Shasthaya batayan (16263); (DGHS) OnHealth24, (DGHS) Mobile Phone Health Service, (DGHS) Telemedicine Service in Union Information & Service Centers, (DGHS) Pregnancy Care Advice through SMS, (Grameenphone)Tonic, (Banglalink) Healthlink, (Bnglalink) Mindcare, (Banglalink) Daktarbhari, (Airtel) Maya Apa, (Airtel) MindTale, (Robi) My Health Family Pack, (Robi) Myhealth Combo, (Teletalk) Shashtho Sheba (Health Care), Doctorsbd, Aponjo
**Kenya** (n=9, 6 fee, 3 free): mDaktari, SIMWAY, myDAWA, Mobile for reproductive health (m4RH), Ilara health, MedAfrica, Hello Doctor/Sema Doc, Daktari popote, MEDBIT
**Nigeria** (n=17, 14 fee, 3 free): Prive Doc, Komplete Care, Hudibia, iCliniq, HealthTap, Tremendoc, MobiHealth, MedAfrica, HealthConnect, DOCS, Reliance Care. Omomi, Seekmed, DoctorNow, NovaDoc, Babylon, Ada

#### Service fees and costs of provider platforms

All but one of the identified services required users to have available airtime or network connectivity. The all-female health provider service, Sehat Kahani (Pakistan) https://sehatkahani.com/, while available through an app, also offers video/phone consultation for patients facilitated by a health worker, from one of 26 physical clinics (none in the study site). Most donor and state-supported services were free at point of use and accounted for 3 of 5 services in Tanzania but only 3/17 in Nigeria and 2/17 in Bangladesh. In Bangladesh, some of the state-provided services charged user fees. In all of the countries, private-for-profit commercial services required users to pay per consultation or via annual membership fees (often with different packages). Some differentiated fees by the user’s health insurance status. Consultation fees ranged widely within and between the countries: US$ 2-30 (Pakistan), US$9-130 (Tanzania), US$5-30 (Kenya), US$0.42-166.67 (Nigeria), US$0.02per day-6per month (Bangladesh). Some services included free follow-up consultations, usually over a specified timeframe. For all, costs were incurred beyond the consultation fee, including any treatment (e.g., drugs) or specialised services on referral. On the supply-side, some services charged membership fees to health workers to belong to the service.

### Use and perceptions of mConsulting platforms

In Bangladesh, Pakistan and Tanzania, health workers and decision-makers recalled services that were no longer operational, mostly due to financial challenges, including withdrawal of donor funding:

> *The funding stopped and then it was handled over to the government! Its main challenge was on payment, because it needs the use of the airtime* […] *something which can be supported by the mobile service companies through the social responsibility* (*Semi-structured interview, mConsulting service provider 1, Tanzania*)

In Bangladesh, some residents mentioned that they had previously used now-defunct services. In addition, some said they were aware of and would or had used currently available government-run platform services, especially during a health emergency.

In the other sites, most community members were unaware of existing platform services and had not used them. Yet, despite little or no direct experience of mConsulting platforms, these were generally perceived to be affordable, especially when free (i.e. only costs of airtime for call/text/internet).

Alongside potential savings on consultation fees, mConsulting was seen to save time and transport costs in congested urban spaces, where traffic is ‘unimaginable’ (Semi-structured interview, Community decision-maker 2, Bangladesh) and in rural areas, where facilities are few and far between:

[…]*access to healthcare services at the right time, so it saves time and costs. Instead of deciding to hire the transport somebody can talk to the doctor at a distance, that is very nice, better than misusing the time through travelling long distance, and you know as much as you delay to access the doctor, the condition continues to be get worse* (*Mini-interview, Community resident 1,Tanzania*).

Other perceived benefits included accessibility to healthcare during local disruptions (e.g., gang activity), industrial action by heath workers in local services, natural disasters (e.g., flooding) or, we add, pandemics. Anonymous consulting for sensitive or stigmatized conditions was seen as an attraction:

> *So if you have gonorrhoea you fear telling* [*your HCW*] *because maybe this is somebody you respect* (*laughing*). *You know illness, illness is a person’s secret. Let’s say maybe I am HIV+, you fear telling her/him* (*Community workshop 1, resident 1, Kenya*).

In Kenya, a few potential users identified the benefit of having direct access to an appropriate health worker, cutting out onerous steps in the care pathway and adding to transparency in the consultation itself. However, some concern was expressed by community residents in Bangladesh that, if of poor quality or inappropriately fitted to the local context, mConsulting would simply add another layer of expense and time, with people having to revert to face-to-face care anyway.

> *They will have to keep in mind about the people they are talking to, and refer accordingly. If they refer to big private hospitals, it will be a problem* (*Semi-structured interview, Community decision-maker 1, Bangladesh*).

Low literacy levels and lack of confidence to use technology were identified as possible barriers to community use of mConsulting platforms in all sites.

Competition with nearby face-to-face services was raised as another possible barrier to using mConsulting platforms in urban Bangladesh:

> *People might think, if they can just go to the pharmacy and get advice and medicine in two minutes, why will they call* [*phone*] *someone they do not know?* (*Community workshop 2, healthcare provider 1, Bangladesh*).

#### (b) Locally available mConsulting with individual health workers using their own mobile phones

In each site, community members and health workers identified examples of mConsulting taking place between residents and locally-based health workers using their own phones, especially during emergencies and after-hours:

> *Sometimes I receive calls informing me ‘Doctor! My son is vomiting’ then I give them emergency advice by telling them to give him/her some water as well as instructing them to take him/her to the nearest health care center* (*Key informant interview, participant 5, Tanzania*).

> *One midnight, my neighbour who is a health provider, was called on the phone for* [*a diarrhoeal*] *complaint. She asked* [*the family*] *not to use* [*ashes and explained*] *that bottled water, salt and sugar should be used, then referred them to any nearby hospital. On getting to the hospital, it was locked. The gateman said, “no doctor nor nurse is available”. The family called the* [*health provider*] *again. She asked if the sick person was still stooling. They said it had subsided* […] *She asked them to keep giving the* [*rehydration*] *solution* […] *I see that the phone was used to communicate* […] *So, in my own view, treatment on the phone is still good* […*if the patient*] *gives the right complaints to who they are complaining to* (*Community workshop 2, resident 1, Nigeria*).

In Bangladesh, individual private-for-profit pharmacists (usually themselves community residents) were usually seen as the first point of phone contact, for community members with queries about minor illness and in emergencies. Pharmacists described listening to symptoms, prescribing and selling treatment, and sometimes making referrals to doctors or hospitals. No fees were charged for these mConsultations, only for drugs sold.

In all of the sites, community leaders, NGO and government health workers similarly mentioned being on call for medical advice and referrals:

> *maybe somebody was sick and wanted to actually maybe inquire the type of drugs that she/he might use in such a condition, instead of going to the hospital spending a lot or even like the matter is urgent as at then, so they inquire from us first before they opt for going to the facility for more treatment* (*Semi-structured interview, healthcare worker 1, Kenya*).

Remote consultations were described between patients and local health workers in cases of care follow-up both for a singular health event (e.g., a medical procedure, acute illness) and for long-term health conditions, such as diabetes and hypertension:

> *My friend got operated on and something was wrong with his stitches. The doctor was not available in town so he called him on the phone and he prescribed the medicine* ((*Semi-structured interview, healthcare worker 3, Pakistan*).

> *Currently we do use the mobile to get in touch with our clients but in a smaller way. Like we do follow-ups using the phone. You know for some reasons there are some clients who may not talk physically* [*face-to-face*] *but if you call them they are able to give you some more information on the phone* (*Semi-structured interview, community-decision maker 1, Kenya*).

### Challenges of mConsulting

While health workers described using their phones to consult with patients in all sites, they did not necessarily recognise their practices as ‘doing mConsulting’. For many, the idea of formally undertaking mConsulting provoked anxiety and apprehension. They felt unconfident about the rules and regulations and wondered how to do mConsulting in a professional and ethical way, if at all:

> *Actually, according to the medical ethics*, [*mConsulting*] *is not allowed except in some few health services, especially in chronic disease, you can usually be asking on the patient’s health progress and provide the advice distantly, you can tell him/her ‘you should be drinking large amounts of water’. But otherwise you should meet with the patient physically* (*Semi-structured interview, healthcare worker 5, Tanzania*).

Some health workers were unconfident about the use of technology. In all study sites, stakeholders raised concerns about data protection and privacy when communicating in a digital world:

> *Aaah, of course it is very difficult to maintain confidentiality when the service has been offered through the mobile communication, as you know the mobile communication passes through different network systems something which creates difficulties in maintaining confidentiality* (*Semi-structured interview, healthcare worker 8, Tanzania*)

Some policy stakeholders flagged practical challenges of keeping up with a rapidly evolving field:

[…] *innovation especially when it’s very rapid, and especially now in technology, is moving light years ahead of the capacity of regulators to stay up to date* (*Key informant interview, participant 2, Nigeria*).

Community members, in Bangladesh, raised concerns about fraudulent service providers operating in the absence of service accreditation or official endorsement. One resident explained that she had been drawn into a fraudulent money transfer scam for ‘hospital services’ (Semi Structured interview, community decision-maker 2). Others mentioned a broader culture of mistrust in digital transaction services, borne from a decade of experience with disreputable online services, particularly money transaction apps, which are prone to fraud.

Across the sites, community members expressed that mConsulting could not be a complete replacement for face-to-face care:

[…] *you know the physical observation assists the doctor in identifying the exact problem facing the client something which results to the appropriate treatment. How will the doctor identify patient’s body temperature and other diagnosis through mConsulting?* (*Mini-interview, community resident 7, Tanzania*)

> *There is a thing of trust in doing anything face-to-face; that I can see you. Now if I call in the dark, who is answering it or not? Therefore this becomes a thing of disbelief. That’s it. We can’t see who was behind the curtain. But if he was face-to-face then we can see that*. (*Community workshop 1, participant 2, Bangladesh*).

Having an opportunity for in-person examination was valued by some policy makers, as well as community members:

> *You cannot compare it* [*mConsulting*] *to when you see patient, for instance now, if* […] *I’m consulting with somebody* [*via mconsulting*] *now; I cannot check how pale the person is*. […] *Even if you do a video, if I say let me see your tongue, the way I’d see it in a video might be different from when I see it* [*physically*] (*Key informant interview, participant 2, Nigeria*)

In Pakistan, some health workers were concerned that mConsulting would negatively impact on the demand for face-to-face services and with this, their livelihoods.

### Finding solutions

To deal with uncertainty and lack of confidence, many local health workers requested specific guidelines and training in the regulations and in the use of technology itself.

### Health workers as intermediaries

In Pakistan, Tanzania, Nigeria and Bangladesh, health workers described using their phones to contact colleagues for advice about patients. Where patients were not present in the conversation, we do not consider this mConsulting. However, a few health workers mentioned occasions when they had consulted with a colleague while with a patient, thereby taking on an intermediary role. In Bangladesh, one pharmacist explained that he had subscribed to a doctor’s app, in order to give medically-sound advice to customers (community workshop 1, healthcare provider 10), effectively serving as a local intermediary for a national mConsulting platform (even although he later unsubscribed due to confusing, expensive service fees charged by the platform). In Tanzania, a community member recalled the benefits of doctors communicating between hospitals to diagnose and prescribe drugs for a newly-admitted patient:

> *So far I remember when my relative was admitted, the doctor spoke to his fellow doctor from* [*Name*] *Hospital through the mobile phone, and actually it was after having done the diagnosis, the doctor gave the instruction to his fellow doctor through mobile phone, then soon we were prescribed drugs* (*Mini-interview, community resident 1, Tanzania*).

### Households as intermediaries

While apprehensions were raised by some community members about the technology and literacy skills needed for mConsulting, some suggested that these are not individual capabilities but distributed household assets - to be shared and translated:

> *Many people are just answering phone calls, they either do not have the time to, or cannot read those text messages. But like I said earlier, at least one member* [*of the family will*] *have a smart phone, so they can teach others. If I am experienced and I have the required knowledge, then I will share that with my family and people around me. For example, if my mother is sick, and I know about mConsulting services, I will call for her* (*Community workshop 2, resident 3, Bangladesh*).

### Integration of mConsulting into existing systems

In Pakistan, the project advisory group, which included experts in digital health and community members and local health workers, suggested development of a ‘hub and spoke model’ to integrate mConsulting within local health infrastructure and as part of a broader telemedicine approach. They suggested training health workers and community members to raise awareness and build capacity for telemedicine, with lady health volunteers working as coordinators. In this model, staff based at primary healthcare facilities would provide referrals for patients to connect remotely to participating secondary and tertiary facilities. The group advised that tele-devices (e.g., for basic laboratory testing) and apps (e.g., for maternal and child health) could be incorporated into this approach too.

In Bangladesh, residents suggested that service accreditation by a trusted stakeholder (e.g., the state or a respected commercial telecommunications company), plus transparency about service details and costs might go some way towards improving trust in digital health services.

> *If they want people to trust, the services should be* [*delivered*] *under the Government. It will be free that’s for sure, but at the end of the month even if they cost something like 20 BDT or 50 BDT, people will accept it*. (*Community workshop 1, healthcare worker 10, Bangladesh*).

They suggested the need for coordination of multiple, fragmented mConsulting services, including those provided by the state, where there is not a common front-end for users:

> *If we become aware that the service is being offered by* […] *all the operators being united and the Government is also involved, more trust and confidence would be built*. (*Community workshop 2, resident 1, Bangladesh*).

## Discussion

Our findings suggest that mConsulting is a viable option for remote and spatially-marginalised communities with minimal access to healthcare services in LMIC settings. In the five countries studied, regulatory frameworks are in place through national ICT and e/mHealth policies^36, 46^-^55, 78^ and mConsulting is already happening: nationally, services delivered through provider platforms are available and at community-level, a few healthcare workers and community members reported direct experience of locally-conducted mConsulting (with healthcare workers using their own phones) - in emergencies, for advice and for care follow-up. Most stakeholders expressed enthusiasm for mConsulting. Much of the mConsulting that we have documented is happening organically from the ground-up. It is not intervention-led and has not been formally evaluated or written up for publication. This has also been noted elsewhere^91, 92^ and may be one reason why we only found a limited number of articles pertaining to mConsulting in the published literature since 2018, despite a growing body of evidence on mobile health more generally in LMIC settings^93^.

While less attention has been given to mConsulting in LMIC settings, our findings resonate with the evidence that we found in our literature review, namely that mConsulting can help to overcome affordability barriers^26^ and facilitate care-seeking practices^20, 24, 27^. Participants identified benefits of convenience and affordability from reduced travel. Consultation fees for services delivered through provider platforms varied between services and across countries; however, free or subsidised services (provided by the state/development partners) were generally perceived to be affordable by potential users (i.e. the cost of airtime for a call). No mention was made of consultation fees for locally-based services (where health workers used their own phones), other than consultations conducted freely by individual private-for-profit pharmacists in Bangladesh who only charged for drugs sold. Consultations initiated by individuals known to each other (as with most of the locally-available consultations described) suggest a form of personalised^26^ and locally-relevant care. mConsulting in this everyday ‘smaller way’ (decision-maker, Kenya), brings opportunity for patients to contact known, trusted healthcare workers and providers to reach patients, especially those who might be harder to reach in person^29^.

Overall, our findings suggest that there is a general willingness amongst decision-makers, healthcare workers and community members to deliver and use mobile consulting services, provided key challenges are addressed. These challenges include tackling the pragmatics of doing mConsulting - technology, infrastructure, data security, confidentiality, acceptability - and ensuring that mConsulting is integrated into wider health and technology systems that are themselves in need of strengthening and support. We identified similar concerns about confidentiality^27^ and continuity of care^29^ in our literature review, along with the potential for mConsulting to reinforce inequalities in patient access to healthcare^19^. A few study participants suggested that lack of technological and literacy skills might impede mConsulting for individuals but suggested these could be overcome by intermediaries. While present in all of the study countries, gender and residential (urban-rural) gaps in mobile phone ownership, important for health access and outcomes^5^, were not explicitly mentioned. However, poor or erratic internet connectivity, unreliable electricity and infrastructural issues were identified as barriers to the implementation of mConsulting in both of the rural settings and regular access to data/WiFi and airtime were issues for residents surveyed in the urban sites.

Our findings are consistent with challenges documented for mHealth more generally in LMIC settings, *viz* uneven/poor network connectivity, rapid technological change, low technological literacy levels amongst users, and limited awareness of available services^3, 94, 95^. Difficulties in integrating health information systems and building mHealth capacity have also been reported^3, 95^ alongside unsupportive policy environments and limited mHealth stewardship, although this is changing, as a growing number of WHO Member States, including those in our study, adopt national digital and eHealth policies^3^. COVID-19 has further forced rapid changes to the policy environment and healthcare delivery^1^. Telemedicine is being promoted in Tanzania’s guidance on mental health and HIV/AIDS services^96^, there has been a surge in online health services in Bangladesh^97^ and in Pakistan, the state has established a COVID-19 Health Advisory Platform^98^ alongside the national telehealth platform.

Pre-COVID, one of the challenges associated with remote consultation was that patients and healthcare workers preferred face-to-face consultations. However, physical distance has become an advantage during the pandemic. The COVID-19 response has prompted reverence for mConsulting rather than it being considered something health workers did on the side, or something occasional for some patients. Dugal et al. (2018) note the importance of a period of adjustment for users and healthcare workers to adapt to using a remote service^27^. By forcing a sudden (and positively-motivated) switch from face-to-face care to mConsulting, COVID-19 has perhaps compressed this period, not only for individual adopters but in society more generally. The level of disruption experienced with COVID-19 may lead to permanent change, with health workers making more systematic use of mConsulting. We can make choices about the nature of that change^99^ to ensure it is undertaken transparently and equitably (for patients and health workers), and use it as opportunity to move towards universal health coverage. How this is, or could be, achieved in terms of policy is likely to vary between countries. This is because the nature of the health system itself, that is, the existing ways in which care is arranged and delivered, its histories and previous experiences with mConsulting and digital services more generally (important to acceptability as noted by study participants in Bangladesh), will be an important determinant of system change^7^. However, at the level of health workers and patients, there may be commonality across health systems on fundamental issues for the delivery of quality healthcare, such as provision of competent care, positive user experience and trust between patient and health provider.^100^

### Strengths and Limitations

Our methodological approach, grounded in complexity theory and systems’ thinking^7, 8^, has afforded comparative insights into multiple perspectives (people, organisations, policies) within and across mConsulting systems. Located in five urban slums and two remote rural contexts in five LMIC countries, our study presents diversity of context, increasing the transferability of our findings to similar low-resource settings. Using a mixed method study design has enabled us to identify the extent of mConsulting within the urban slum sites and to understand reasons for this. Furthermore, we have been able to draw in lessons from our review of current evidence to contextualise the findings of our scoping study in urban and rural settings, thereby providing a more comprehensive picture than would be possible with quantitative or qualitative methods alone^89^.

Many community residents and health workers engaged enthusiastically with the ‘idea’ of mConsulting without first-hand experience, especially of formal provider platforms. We expect that unanticipated challenges and benefits will emerge as mConsulting is further introduced and adopted within low resource settings. However, perceptions and willingness are important drivers of usage intention in telemedicine^101^, as well as of health-seeking behaviour, community trust and service acceptability^10^.

### Implications for policy and practice

Our study findings suggest various policy avenues for strengthening and supporting mConsulting in low-resource communities in LMIC settings, including:

*Equipping and enabling healthcare workers to deliver mConsulting* by:

a. Developing guidance to situate mConsulting as part of professional and ethical conduct (using national, regional and international regulatory frameworks, including COVID-19-related guidance^2^ and planning tools such as the *Digital implementation investment guide*^102^ provided by WHO).
b. Developing guidance for the protection of data and privacy.
c. Training and mentoring health workers to enable a confident transition between face-to-face and audio/text-based consulting, including knowing when to see their patients in person; and managing new ways of working.
d. Equipping health workers with airtime and either providing the hardware (ideally smartphones, rechargeable or solar batteries, especially in contexts where electricity is unreliable) or supporting health worker use of their own phone (e.g., second sim card in countries where two sim phones are common, secure apps for work use, maintenance costs).

*Embedding and integrating mConsulting provider platforms within the wider health system* through:

a. Incorporating them into existing accreditation, regulation and governance structures^102^.
b. Raising awareness of mConsulting by a trusted healthcare provider or leader (nationally and using community structures) to facilitate community understanding and trust in this form of clinical communication.
c. Adapting platform services to local conditions and ensuring appropriate and accessible prescriptions and referrals

*Considering ways to support local health workers to take on a boundary spanning role between national provider platforms and local communities* by assisting them to use their phones in ways that connect them to platform providers as well as community members (as the pharmacist in Bangladesh sought to do). By sharing information, translating knowledge and linking different groups, boundary spanners may facilitate system integration, improve functionality, build trust and bring services closer to those who need them^103, 104^.

*Enabling patients to confidently and effectively use mConsulting services* by raising community knowledge about how mConsulting services work, what happens if an examination, test or referral is needed and how service providers maintain confidentiality and data security; and how patients can ensure confidentiality on their side.

## Conclusion

mConsulting has the potential to strengthen health systems during and beyond the COVID-19 global pandemic. However, a whole system approach is needed, one that recognises mConsulting as one component of the care pathway. While there are indications of local readiness for mConsulting in communities with minimal access to healthcare, wider system strengthening is needed to bolster referral and specialist services, laboratories and supply-chains in order to fully realise the continuity of care and responsiveness that mConsulting services can offer.

## Supporting information

Supplemental file - ethics and reporting

## Data Availability

No additional data are available.

## Declarations

### Funder acknowledgement

This work was supported by a foundation grant from the Health Systems Research Initiative with funding from the UK Department for International Development, the UK Economic and Social Research Council, the UK Medical Research Council and Wellcome (Grant no. MR/S012729/1). SP and MA (Global Challenges Research Fund Fellowship No. IAS/32013/19) gratefully acknowledge support provided by the Warwick Institute of Advanced Study. FG receives funding as South Africa Research Chair in Health Policy and Systems from the National Research Foundation, South Africa. RJL is supported by the NIHR Applied Research Collaboration (ARC) West Midlands, UK.

### Ethical approval

All participants provided informed consent to participate before taking part in the study. Ethical approval was obtained from:

AMREF Health Africa Ethics and Scientific Review Committee (AMREF-ESRC P719/2019) Biomedical and Scientific Research Ethics Sub-Committee, University of Warwick, United Kingdom (REGO-2019-2343)

National Institute for Medical Research, Tanzania (NIMR/HQ/R.8a/Vol.IX/3044) Pakistan: Ethics Review Committee ERC (No: 2019-1040-3484)

Research Ethics Committee of the Oyo State Ministry of Health (AD13/479/1193).

The Institutional Review Board of the Institute of Health Economics (IHE-IRB), which is approved by Federalwide Assurance (FWA), Bangladesh (No. FWA00026031)

We analysed secondary data obtained through the NIHR Global Health Research Unit on Improving Health in Slums. This work was granted full ethical approval by:

AMREF Health Africa Ethics and Scientific Review Committee (AMREFESRC P440/2018) Bangladesh Medical Research Council BMRC/NREC/2016-2019/759).

Biomedical and Scientific Research Ethics Sub-Committee, University of Warwick (REGO-2017-2043 AM01),

Ministry of Health, Lagos State Government (LSMH/2695/11/259), Research Ethics Committee of the Oyo State Ministry of Health, Nigeria (AD13/479/657)

National Bioethics Committee Pakistan (4-87/NBC-298/18/RDC3530)

### Guarantor

FG

### Contributorship

FG conceived and led the mConsulting study. All authors except RP contributed to development of the project concept, study design and research questions presented. BH wrote the first draft of the manuscript; JAW conducted the literature review; MA, PB, BC, NC, OF, PK, NR analysed site-specific data. BH, FG, JS, BC, SP, AK, MA, AO, EO, NR, PK, NC, JAW, RP and TNA developed the manuscript; All authors reviewed and approved the submitted version of the manuscript.

## Acknowledgements

We would like to thank all study participants for generously sharing their experiences and insights. We are grateful to the NIHR Global Health Research Unit on Improving Health in Slums for providing access to the data collected for the household and adult surveys and for providing contextual grounding for our work. Special thanks are extended to Samuel I Watson and Syed A.K. Shifat Ahmed for providing support with the secondary data analysis, and to Samantha Johnson for assisting with the search strategy used in our literature review.

---

**Appendix 1**. Search strategy followed in literature review of current evidence for mConsulting in LMIC contexts

Our search strategy was developed with an academic support librarian and guided by previous reviews led by Griffiths and Sturt on two-way digital clinical communication with patients^12-18^; with added parameters for LMICs. We used MeSH, free-text, sub-heading, truncation (*) and wildcards ($) for the concepts of ‘mobile’ and ‘consulting’.

We searched key databases: Medline, Embase, Web of Science and Google Scholar for reviews and empirical studies on mConsulting in LMICs, published between 01/01/2018-27/04/2020. This timeframe coincides with the emergence of WHO guidance on digital interventions for health systems s^11^ and recognises the rapidly changing nature of mobile and digital technology.

We included reviews and empirical studies from any LMIC setting with intervention(s) that clearly involve two-way digital communication between patient/user and health provider; and any study published in English. Studies from high-income countries only were excluded. Duplicates were removed, titles and abstracts screened and we reviewed a full copy of each included paper. Data were extracted and findings synthesised.

We identified 207 reviews and 62 empirical studies from the combined database searches. Following the screening of titles and abstracts, we included 36 reviews and 28 empirical studies for full text review. After reviewing the full papers, seven reviews^19-25^ and five empirical studies^26-30^ met the inclusion criteria.

**Appendix 2**. Questions analysed in the **adult** and household **[HH]** surveys collected as part of the NIHR Global Health Research Unit on Improving Health in Slums^80, 81^.

|  |
| --- |
| **Q110A.** Does respondent in (q101a) carry at least one mobile phone day-to-day? **[HH]** |
| **Q110B.** Does the respondent in (q101a) have access to and is the respondent in (q101a) able to use a computer, tablet or other form of digital communication other than a mobile phone, day-to-day?**[HH]** |
| **Q223.** How many days in the month do you have air time (for calls and SMS) for at least one mobile phone in the household **[HH]** |
| **Q224.** How many days in the month does someone in the household have data or access to wifi (for accessing internet for searching the web, using social media or using Email) for at least one of your digital communication devices in the household (smart phone, laptop, tablet)? **[HH]** |
| **Q435** In the last 12 months have you used or attempted to use your mobile phone or other digital communication device (e.g. laptop, tablet) to access health information, advice or care for yourself, where information about your health was received or given?**[adult]** |

