## Supplemental file - ethics and reporting for "Mobile consulting (mConsulting) as an option for accessing healthcare services for communities in remote rural areas and urban slums in low- and middle- income countries: A mixed methods study"

**Supplementary Material: Ethics and Reporting (using guidance and checklists as suggested by the EQUATOR network )**
Mobile consulting (mConsulting) as an option for accessing healthcare services for communities in remote rural areas and urban slums in low- and middle- income countries: A mixed methods study

Our paper is a mixed methods study, drawing on data from:
**1. Current evidence for mobile (m)Consulting through a review of the literature**. To place our study within the context of current evidence, we followed a systematic process of searching and identifying literature, guided by key elements of the PRISMA approach (see checklist below). We have used the results from our search to provide background and context to our study. Due to space constraints within our paper, further details of our approach are available on request.

**2. Secondary data analysis from household and adult survey data (NIHR Global Health Research Unit on Improving Health in Slums)**
Details of the primary study are available: Bakibinga, P., et al., *A protocol for a multi-site, spatially-referenced household survey in slum settings: methods for access, sampling frame construction, sampling, and field data collection.* BMC Medical Research Methodology, 2019. **19**(1) (noted in the paper) and on request from the study team. For our analysis, we tabulated data from the relevant sections of the household and adult surveys (mobile phone, internet, airtime access, use of technology for health-care seeking). We have included these questions in Appendix 2. For each site, the total sample for that site was tabulated against the total number of respondents for that particular question per site. This is explained on p.16. The results are presented (pp.17-18) and integrated in discussion/recommendations to inform policy response (pp.24-27)

**3. Qualitative interviews and workshops with key stakeholders** - reported using Standards for Reporting Qualitative Research (SRQR), EQUATOR Network research reporting checklist: mConsulting study, below). Further information is available on request.

**Ethical approval**
All participants provided informed consent to participate before taking part in the study. Ethical approval was obtained from:
AMREF Health Africa Ethics and Scientific Review Committee (AMREF-ESRC P719/2019)
Biomedical and Scientific Research Ethics Sub-Committee, University of Warwick, United Kingdom (REGO-2019-2343)
National Institute for Medical Research, Tanzania (NIMR/HQ/R.8a/Vol.IX/3044)
Pakistan: Ethics Review Committee ERC (No: 2019-1040-3484)
Research Ethics Committee of the Oyo State Ministry of Health (AD13/479/1193).
The Institutional Review Board of the Institute of Health Economics (IHE-IRB), which is approved by Federalwide Assurance (FWA), Bangladesh (No. FWA00026031)

**PRISMA 2009 Checklist,** Moher D, Liberati A, Tetzlaff J, Altman DG, The PRISMA Group (2009). Preferred Reporting Items for Systematic Reviews and Meta-Analyses: The PRISMA Statement. PLoS Med 6(7): e1000097. doi:10.1371/journal.pmed1000097 [www.prisma-statement.org](www.prisma-statement.org).

| Section/topic | # | Checklist item | Reported on page # |
|---|---|---|---|
| **TITLE** | | | |
| Title | 1 | Identify the report as a systematic review, meta-analysis, or both. | N/A |
| **ABSTRACT** | | | |
| Structured summary | 2 | Provide a structured summary including, as applicable: background; objectives; data sources; study eligibility criteria, participants, and interventions; study appraisal and synthesis methods; results; limitations; conclusions and implications of key findings; systematic review registration number. | N/A |
| **INTRODUCTION** | | | |
| Rationale | 3 | Describe the rationale for the review in the context of what is already known. | pp.5-6 |
| Objectives | 4 | Provide an explicit statement of questions being addressed with reference to participants, interventions, comparisons, outcomes, and study design (PICOS). | PICOS N/A |
| **METHODS** | | | |
| Protocol and registration | 5 | Indicate if a review protocol exists, if and where it can be accessed (e.g., Web address), and, if available, provide registration information including registration number. | On request, registration N/A |
| Eligibility criteria | 6 | Specify study characteristics (e.g., PICOS, length of follow-up) and report characteristics (e.g., years considered, language, publication status) used as criteria for eligibility, giving rationale. | Appendix 1, p.36 |
| Information sources | 7 | Describe all information sources (e.g., databases with dates of coverage, contact with study authors to identify additional studies) in the search and date last searched. | |
| Search | 8 | Present full electronic search strategy for at least one database, including any limits used, such that it could be repeated. | On request |
| Study selection | 9 | State the process for selecting studies (i.e., screening, eligibility, included in systematic review, and, if applicable, included in the meta-analysis). | Appendix 1, p.36 and on request |
| Data collection process | 10 | Describe method of data extraction from reports (e.g., piloted forms, independently, in duplicate) and any processes for obtaining and confirming data from investigators. | On request |
| Data items | 11 | List and define all variables for which data were sought (e.g., PICOS, funding sources) and any assumptions and simplifications made. | |
| Risk of bias in individual studies | 12 | Describe methods used for assessing risk of bias of individual studies (including specification of whether this was done at the study or outcome level), and how this information is to be used in any data synthesis. | |
| Summary measures | 13 | State the principal summary measures (e.g., risk ratio, difference in means). | |
| Synthesis of results | 14 | Describe the methods of handling data and combining results of studies, if done, including measures of consistency (e.g., $I^2$) for each meta-analysis. | |

| Section/topic | # | Checklist item | Reported on page # |
|---|---|---|---|
| Risk of bias across studies | 15 | Specify any assessment of risk of bias that may affect the cumulative evidence (e.g., publication bias, selective reporting within studies). | N/A |
| Additional analyses | 16 | Describe methods of additional analyses (e.g., sensitivity or subgroup analyses, meta-regression), if done, indicating which were pre-specified. | N/A |
| **RESULTS** | | | |
| Study selection | 17 | Give numbers of studies screened, assessed for eligibility, and included in the review, with reasons for exclusions at each stage, ideally with a flow diagram. | p.5, flow diagram on request |
| Study characteristics | 18 | For each study, present characteristics for which data were extracted (e.g., study size, PICOS, follow-up period) and provide the citations. | On request |
| Risk of bias within studies | 19 | Present data on risk of bias of each study and, if available, any outcome level assessment (see item 12). | N/A |
| Results of individual studies | 20 | For all outcomes considered (benefits or harms), present, for each study: (a) simple summary data for each intervention group (b) effect estimates and confidence intervals, ideally with a forest plot. | On request |
| Synthesis of results | 21 | Present results of each meta-analysis done, including confidence intervals and measures of consistency. | N/A |
| Risk of bias across studies | 22 | Present results of any assessment of risk of bias across studies (see Item 15). | N/A |
| Additional analysis | 23 | Give results of additional analyses, if done (e.g., sensitivity or subgroup analyses, meta-regression [see Item 16]). | N/A |
| **DISCUSSION** | | | |
| Summary of evidence | 24 | Summarize the main findings including the strength of evidence for each main outcome; consider their relevance to key groups (e.g., healthcare providers, users, and policy makers). | p.24-27 |
| Limitations | 25 | Discuss limitations at study and outcome level (e.g., risk of bias), and at review-level (e.g., incomplete retrieval of identified research, reporting bias). | |
| Conclusions | 26 | Provide a general interpretation of the results in the context of other evidence, and implications for future research. | |
| **FUNDING** | | | |
| Funding | 27 | Describe sources of funding for the systematic review and other support (e.g., supply of data); role of funders for the systematic review. | p. 26 |

**Standards for Reporting Qualitative Research (SRQR), EQUATOR Network research reporting checklist: mConsulting study**

O'Brien, Bridget C.; Harris, Ilene B.; Beckman, Thomas J.; Reed, Darcy A.; Cook, David A. Standards for Reporting Qualitative Research: A Synthesis of Recommendations. Academic Medicine89(9):1245-1251, September 2014. doi: 10.1097/ACM.0000000000000388

| **No** | **Topic (see item description in Table 2 below)** | **Page in manuscript (as collated pdf)** |
|---|---|---|
| **Title and abstract** | | |
| S1 | Title | p. 1 (Mixed methods) |
| S2 | Abstract | p. 3 |
| **Introduction** | | |
| S3 | Problem formulation | p.4 |
| S4 | Purpose of research question | p.4, p.14 |
| **Methods** | | |
| S5-15 | Qualitative approach and research paradigm | p.14 |
| | Researcher characteristics/reflexivity | p.15/on request |
| | Context | pp.7-14 (national and site settings) |
| | Sampling | p. 15, p.28 |
| | Ethical issues | p. 15 and on request |
| | Data collection methods | p. 15 |
| | Data collection instruments | p. 15 |
| | Units of study | p. 15 |
| | Data processing | p. 16 |
| | Data analysis | p. 16 |
| | Techniques to enhance trustworthiness | p. 16 |
| **Results/findings** | | |
| S16 | Synthesis and interpretation | p. 17-24 |
| S17 | Links to empirical data | Through evidence review, sampling (p.15) and integrated in discussion (p.24-27) |
| **Discussion** | | |
| S18 | Integration with prior work, implications, transferability, contributions to the field | p. 24-27 |
| S19 | Limitations | pp. 26-27 |
| **Other** | | |
| S20 | Conflicts of interest | p. 28 None declared |
| S21 | Funding | p. 28 |

### Table 2 - description of items (from O'Brein et al, 2014)

| No. | Topic | Item |
| --- | --- | --- |
| | **Title and abstract** | |
| S1 | Title | Concise description of the nature and topic of the study Identifying the study as qualitative or indicating the approach (e.g., ethnography, grounded theory) or data collection methods (e.g., interview, focus group) is recommended |
| S2 | Abstract | Summary of key elements of the study using the abstract format of the intended publication; typically includes background, purpose, methods, results, and conclusions |
| | **Introduction** | |
| S3 | Problem formulation | Description and significance of the problem/phenomenon studied; review of relevant theory and empirical work; problem statement |
| S4 | Purpose or research question | Purpose of the study and specific objectives or questions |
| | **Methods** | |
| S5 | Qualitative approach and research paradigm | Qualitative approach (e.g., ethnography, grounded theory, case study, phenomenology, narrative research) and guiding theory if appropriate; identifying the research paradigm (e.g., postpositivist, constructivist/ interpretivist) is also recommended; rationale[b] |
| S6 | Researcher characteristics and reflexivity | Researchers' characteristics that may influence the research, including personal attributes, qualifications/experience, relationship with participants, assumptions, and/or presuppositions; potential or actual interaction between researchers' characteristics and the research questions, approach, methods, results, and/or transferability |
| S7 | Context | Setting/site and salient contextual factors; rationale[b] |
| S8 | Sampling strategy | How and why research participants, documents, or events were selected; criteria for deciding when no further sampling was necessary (e.g., sampling saturation); rationale[b] |
| S9 | Ethical issues pertaining to human subjects | Documentation of approval by an appropriate ethics review board and participant consent, or explanation for lack thereof; other confidentiality and data security issues |
| S10 | Data collection methods | Types of data collected; details of data collection procedures including (as appropriate) start and stop dates of data collection and analysis, iterative process, triangulation of sources/methods, and modification of procedures in response to evolving study findings; rationale[b] |
| S11 | Data collection instruments and technologies | Description of instruments (e.g., interview guides, questionnaires) and devices (e.g., audio recorders) used for data collection; if/how the instrument(s) changed over the course of the study |
| S12 | Units of study | Number and relevant characteristics of participants, documents, or events included in the study; level of participation (could be reported in results) |
| S13 | Data processing | Methods for processing data prior to and during analysis, including transcription, data entry, data management and security, verification of data integrity, data coding, and anonymization/deidentification of excerpts |
| S14 | Data analysis | Process by which inferences, themes, etc., were identified and developed, including the researchers involved in data analysis; usually references a specific paradigm or approach; rationale [b] |
| S15 | Techniques to enhance trustworthiness | Techniques to enhance trustworthiness and credibility of data analysis (e.g., member checking, audit trail, triangulation); rationale[b] |
| | **Results/findings** | |
| S16 | Synthesis and interpretation | Main findings (e.g., interpretations, inferences, and themes); might include development of a theory or model, or integration with prior research or theory |
| S17 | Links to empirical data | Evidence (e.g., quotes, field notes, text excerpts, photographs) to substantiate analytic findings |
| | **Discussion** | |
| S18 | Integration with prior work, implications, transferability, and contribution(s) to the field | Short summary of main findings; explanation of how findings and conclusions connect to, support, elaborate on, or challenge conclusions of earlier scholarship; discussion of scope of application/ generalizability; identification of unique contribution(s) to scholarship in a discipline or field |
| S19 | Limitations | Trustworthiness and limitations of findings |
| | **Other** | |
| S20 | Conflicts of interest | Potential sources of influence or perceived influence on study conduct and conclusions; how these were managed |
| S21 | Funding | Sources of funding and other support; role of funders in data collection, interpretation, and reporting |

[a]The authors created the SRQR by searching the literature to identify guidelines, reporting standards, and critical appraisal criteria for qualitative research; reviewing the reference lists of retrieved sources; and contacting experts to gain feedback. The SRQR aims to improve the transparency of all aspects of qualitative research by providing clear standards for reporting qualitative research.

[b]The rationale should briefly discuss the justification for choosing that theory, approach, method, or technique rather than other options available, the assumptions and limitations implicit in those choices, and how those choices influence study conclusions and transferability. As appropriate, the rationale for several items might be discussed together.

ACADEMIC MEDICINE
